# Covid spirals: a phase diagram representation of COVID-19 effective reproduction number *R*_*t*_

**DOI:** 10.1101/2021.09.02.21262861

**Authors:** K. W. Pesenti, R. Pesenti

## Abstract

In this paper, we propose a phase diagram representation of COVID-19 effective reproduction number *R*_*t*_. Specifically, we express *R*_*t*_ as a function of the estimated infected individuals. This function plots a particular clockwise spiral that allows to easily compare the evolution of the number of new infected individuals at different dates and, possibly, provide some hints on the future progression of the infection.

## 1 Background

This study is motivated by a collaboration between the Ca’ Foscari University of Venice, Italy, with the health authorities of the provinces of Triest and Gorizia in the North-Eastern Italian region Friuli Venezia-Giulia.

Specifically, our interest is determining whether the current evolution of the infection is similar to the one previously observed in some specific period of the past year, with the aim of supporting decisions, e.g., about the (re)introduction of containment measures.

In this paper, we show that the representation of the effective reproduction number *R*_*t*_ [3] *as a function of the estimated infected individuals may provide an, at least partial, answer to our questions. The plot of the function R*_*t*_ = *R*_*t*_(*I*_*t*_), being *I*_*t*_ the number of infectious individuals at time *t*, in the space *I × R*_*t*_ describes a clockwise spiral. This spiral allows us to easily compare the evolution of the number of new infected individual at different dates and, possibly, provide also some hints on the future progression of the infection.

The rest of the paper is organized as follows:

- Section 2 presents the methods implemented, also providing the source of the data.
- Section 3 discusses the possible uses of the *R*_*t*_(*I*_*t*_) graph and points out the relations between the *R*_*t*_(*I*_*t*_) graph and the different phases of the COVID-19 infection evolution.
- Section 4 draws some conclusions.

## 2 Methods

Italy is (roughly) organized in constituent entities of three different levels. Regions are the first-level constituent entities. Each region is subdivided in provinces, i.e., the second-level constituent entities. Finally, the municipalities are the lowest constituent entities.

Each day, the Italian Civil Protection collects the number of new recorded infected individuals from the health authorities of each Italian province. Then, the Civil Protection makes these data openly available at https://github.com/pcm-dpc/COVID-19.

In this work, we use the Civil Protection data to estimate both *R*_*t*_ and *I*_*t*_ values. Specifically, we compute these two values using the methodology proposed in [2] for both the regions and the provinces of Italy. Finally, we plot the values assumed by *R*_*t*_ and *I*_*t*_ over time in the space *I × R*_*t*_ to obtain a spiral graph.

Our estimates and the associated graphs are available at http://virgo.unive.it/pesenti/tekwp/dashboard.php. Here, a word of caution must be spent. The data recorded by the health authorities may sometimes present some errors that must be corrected, otherwise a negative number of new infected individuals may be obtained on some days. Our way of correcting these errors may differs from the one by other authors. Then, our estimated values for *R*_*t*_ may present small discrepancies, e.g., from the values estimated by the Italian National Institute for Nuclear Physics at https://covid19.infn.it/.

## 3 Results

In this section, we describe the relations that occur between the graph of the function *R*_*t*_(*I*_*t*_) and the number of the new infected individuals as they emerge from empirical data concerning the spread of the COVID-19 infection in the Italian regions and provinces.

### 3.1 Loops and waves

Figure 1 presents the graph of the values of the *R*_*t*_(*I*_*t*_) function observed in the province of Milan (the second most populated Italian province) between August 25th, 2020 and August 24th, 2021. The graph includes three main clockwise loops which correspond to the three main pandemic waves occurred in Milan (see Fig. 2). The spread of contagion is measured in terms of the 7-day moving average number of new infected individuals per day. We consider a 7-day moving average as a smaller number of potential infected individuals are tested on the week-ends than in the week-days in Italy.

**Figure 1:**
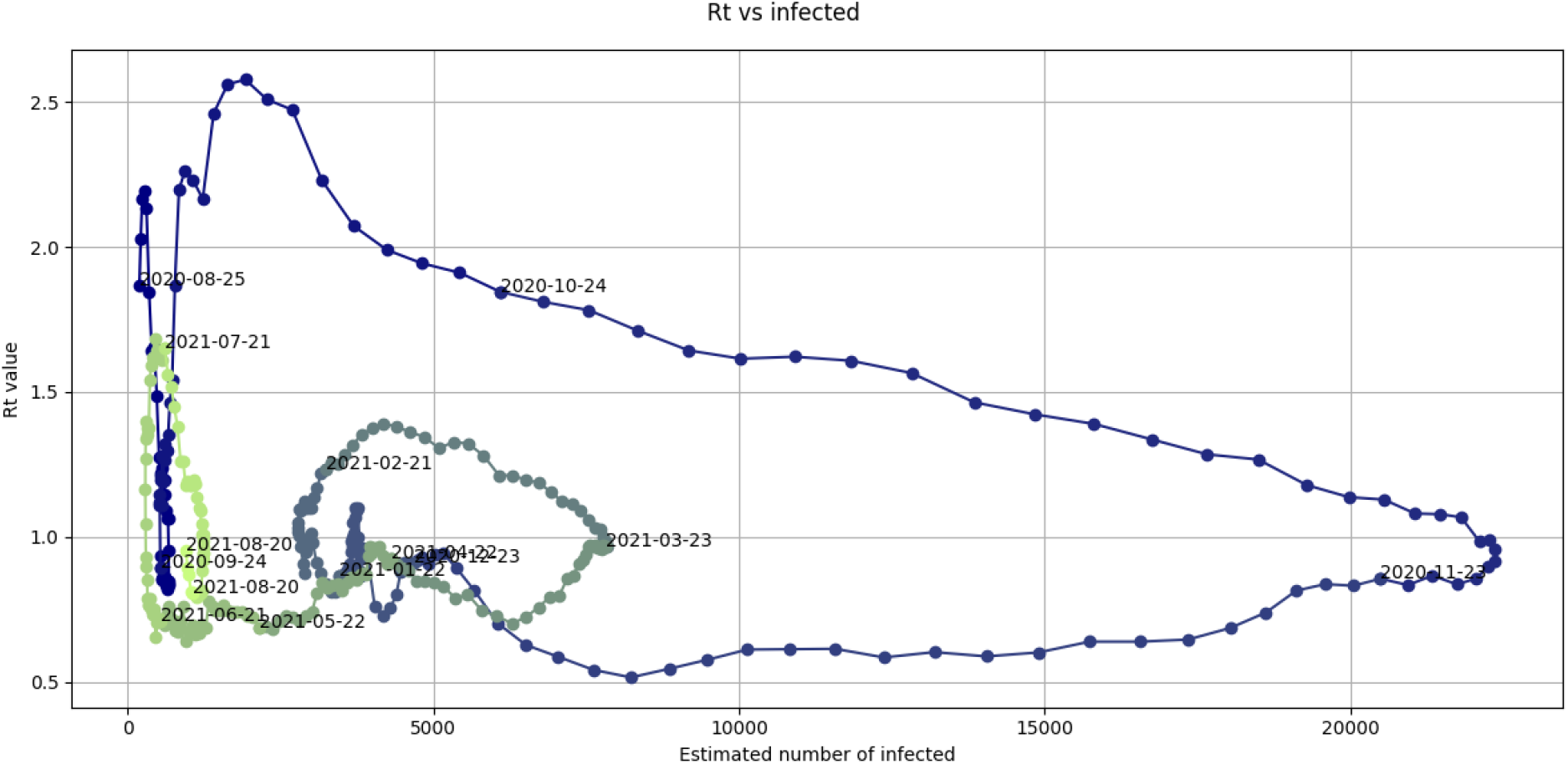
Graph of the values of the *R*_*t*_(*I*_*t*_) function observed in the province of Milan between August 25th, 2020 and August 24th, 2021.

**Figure 2:**
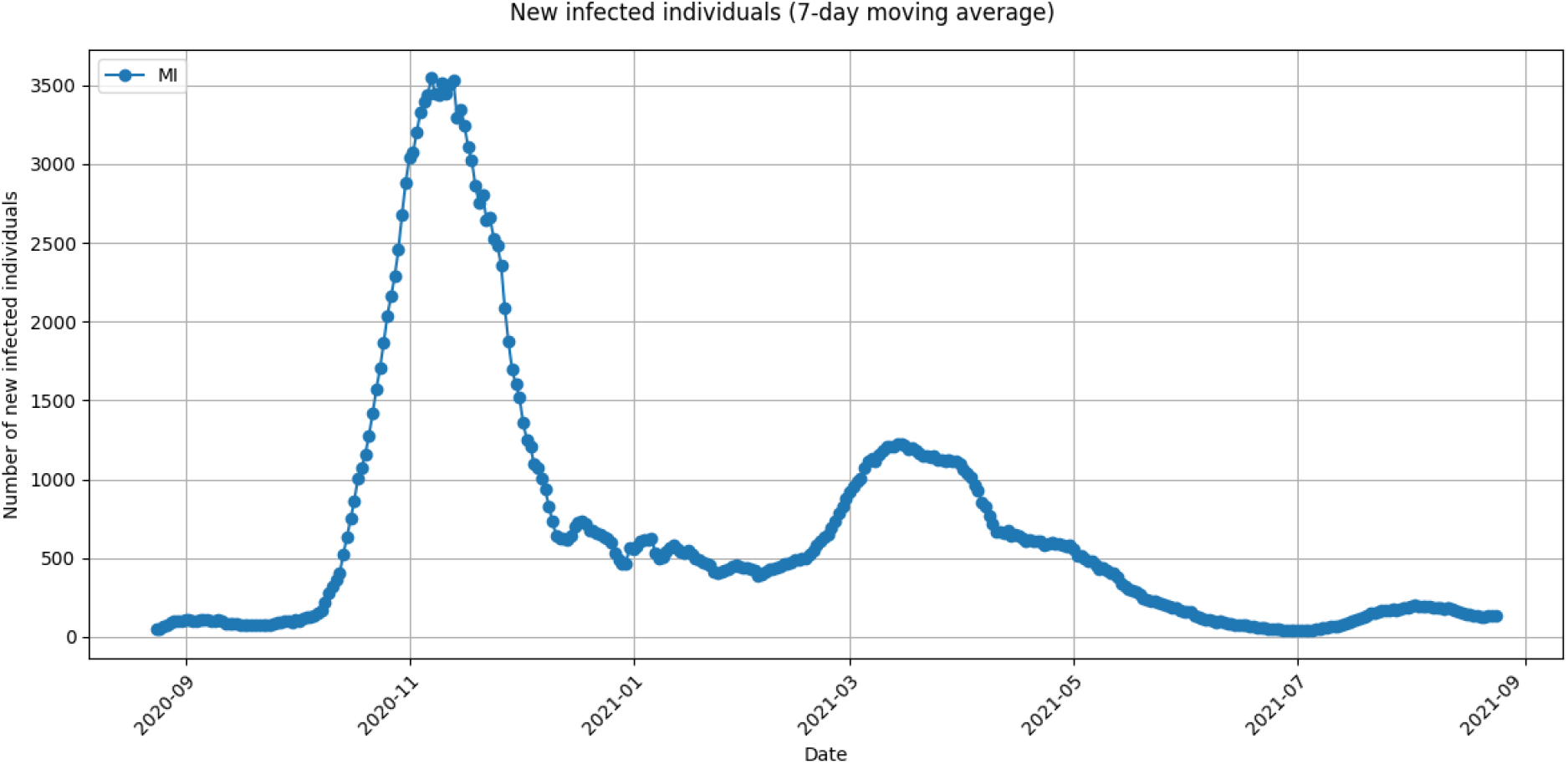
7-day moving average of the number of new infected individuals observed in the province of Milan between August 25th, 2020 and August 24th, 2021.

The loops of the graph in Fig. 1 are represented in different colors. The first and biggest loop is the blue one. It corresponds to the pandemic wave that reached its maximum on November 10th, 2020. The second loop is the gray one. It corresponds to the pandemic wave that reached its maximum on March 14th, 2021. The third wave is the green one. It corresponds to the pandemic wave that reached its maximum on August 2nd, 2021.

Figure 3 is a zoom in of Fig. 1 for the months of July and August, 2021, and must be confronted with Fig. 4, i.e., the zoom in of Fig. 2. In Fig. 3, we can observe a steep increase of the value of *R*_*t*_ in the very first days of July, 2021, being the number of the estimated infected individuals almost constant. This steep increase corresponds to the very beginning of the pandemic wave represented in Fig. 4. Similar almost vertical increases of the value of *R*_*t*_ can be observed in correspondence of the very beginning of the previous two pandemic waves represented in Fig. 2.

**Figure 3:**
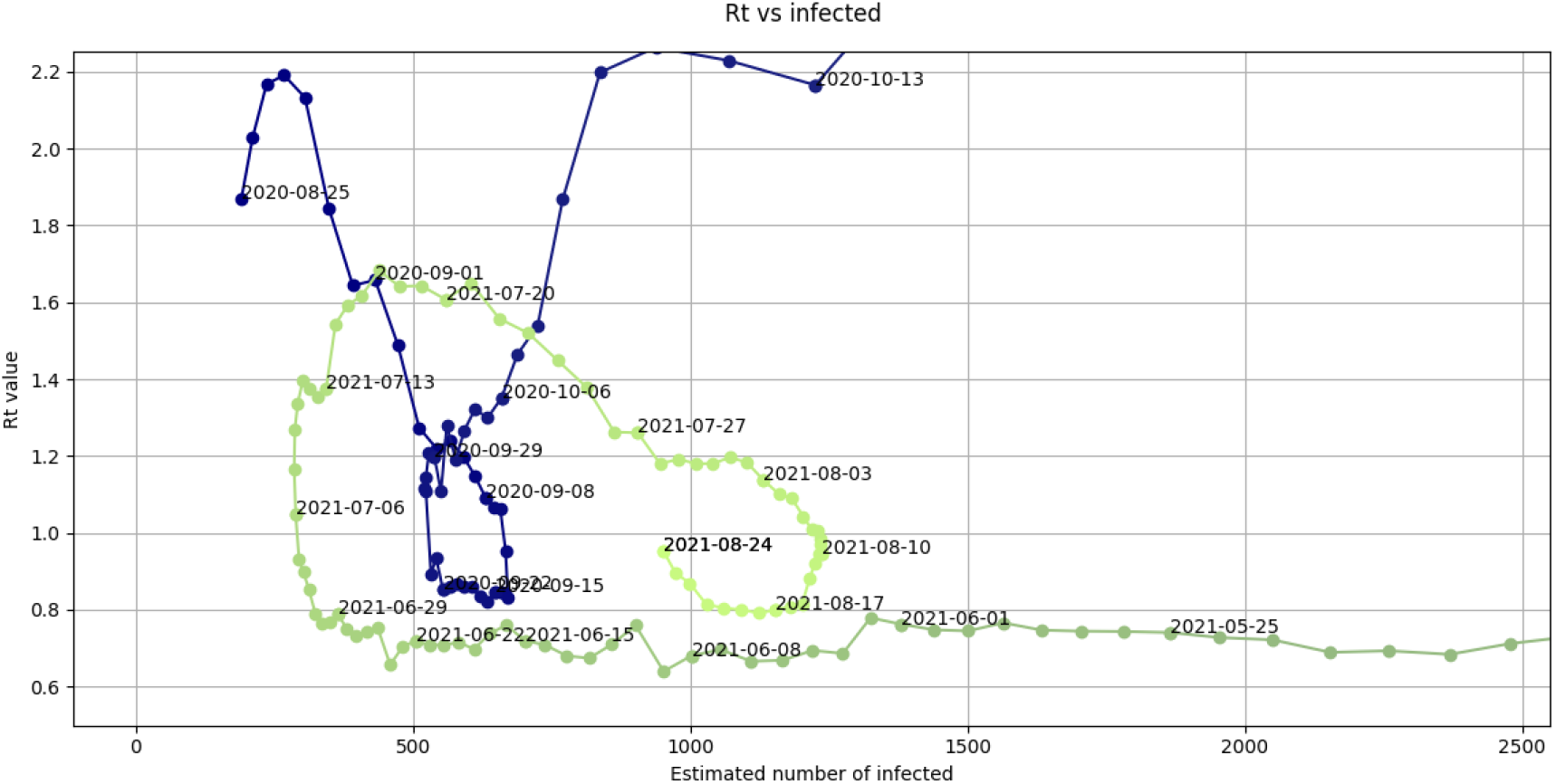
Zoom in of the graph in Fig. 1 where the values of the *R*_*t*_(*I*_*t*_) function in the months of July and August 2021 are highlighted.

**Figure 4:**
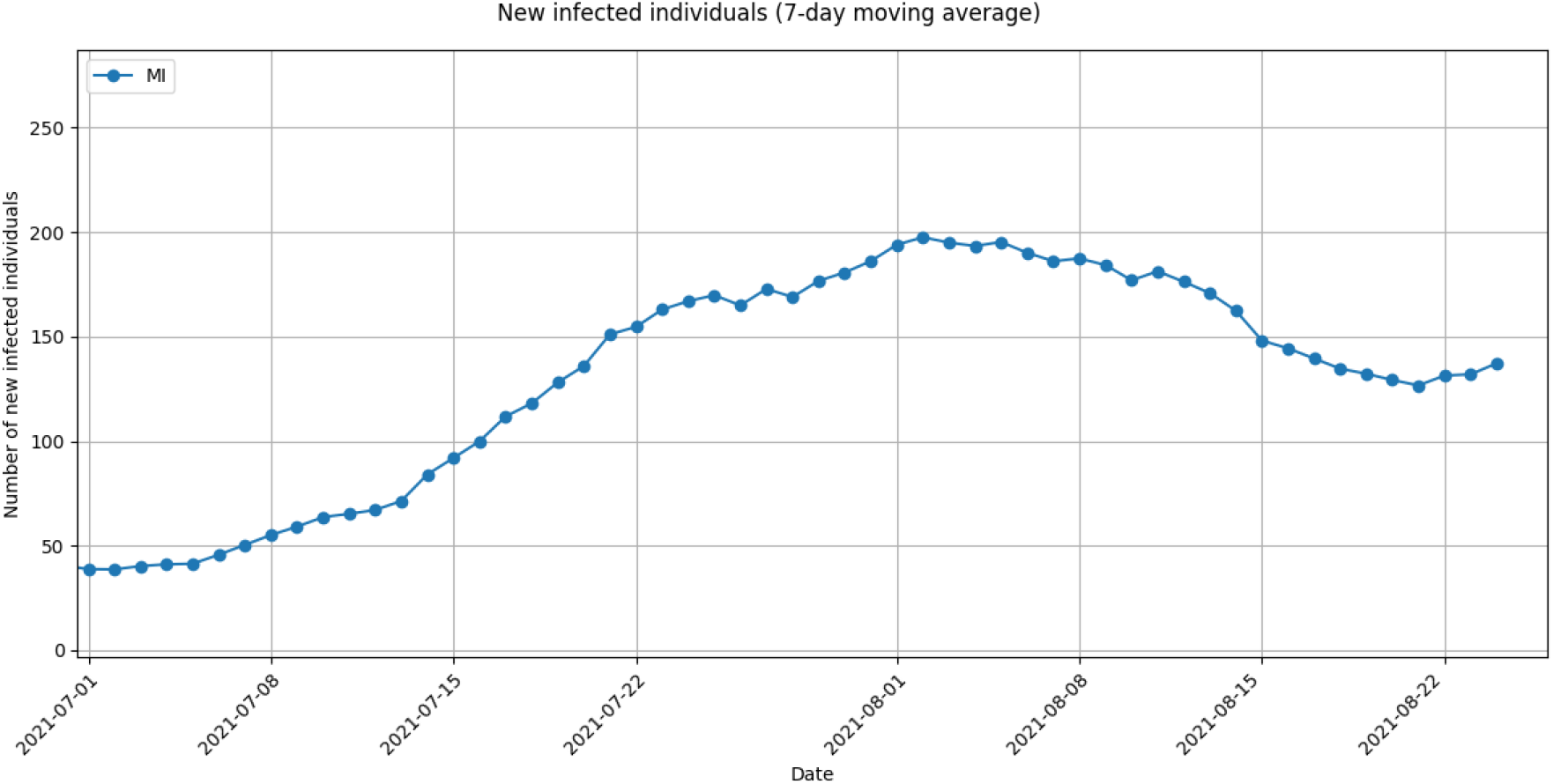
Zoom in of the graph in Fig. 2 for the months of July and August 2021.

Figure 5 and its zoom in (Fig. 6) for the months of July and August, 2020, allow the reader to compare the information that can be deduced from the graph function *R*_*t*_(*I*_*t*_) with the one deriving from the following index:

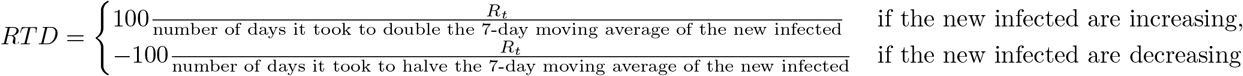

**Figure 5:**
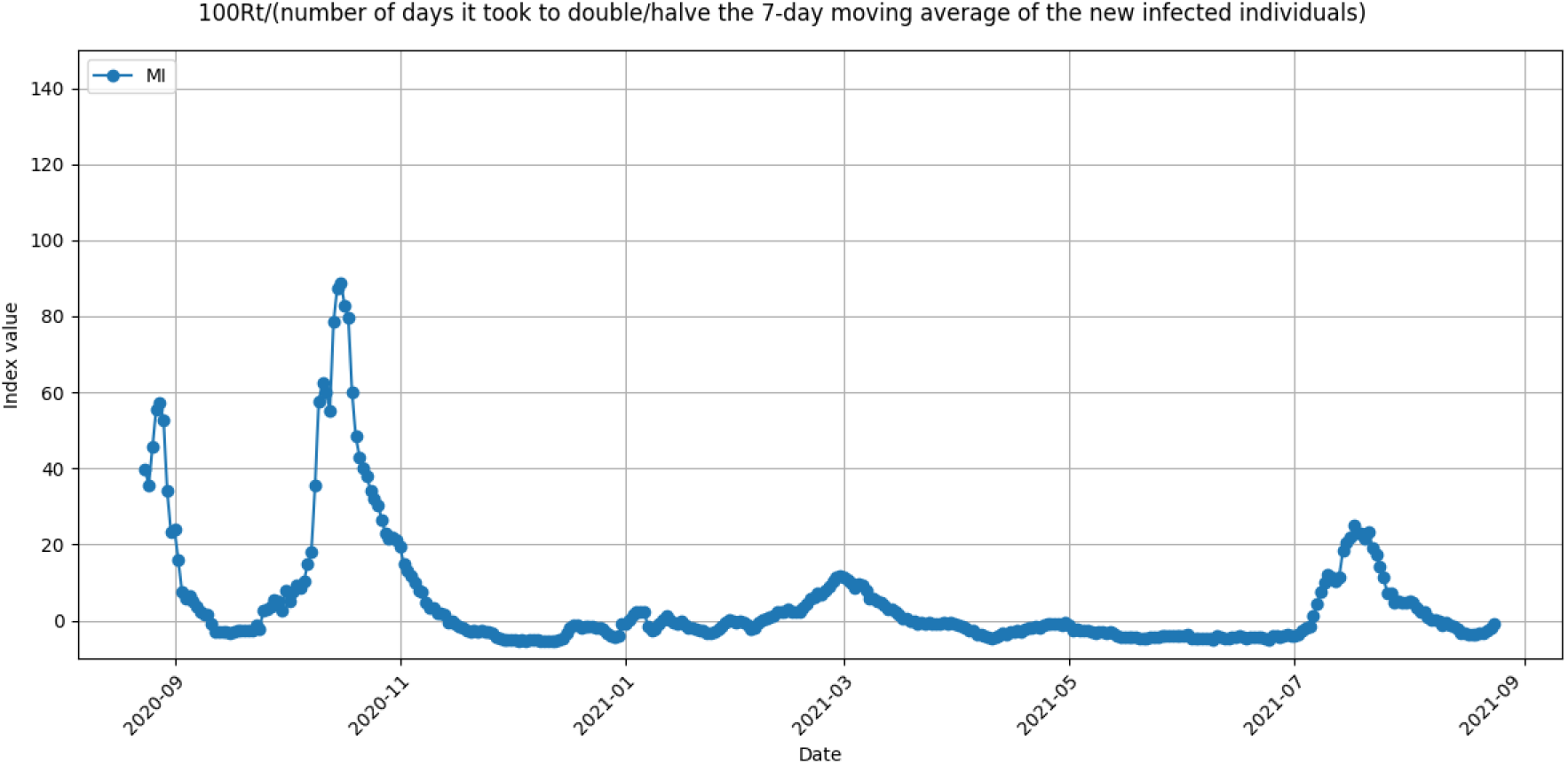
Ratio between *R*_*t*_ and the number of days it took to double/halve the 7-day moving average of the number of new infected individuals observed in the province of Milan between August 25th, 2020 and August 24th, 2021.

**Figure 6:**
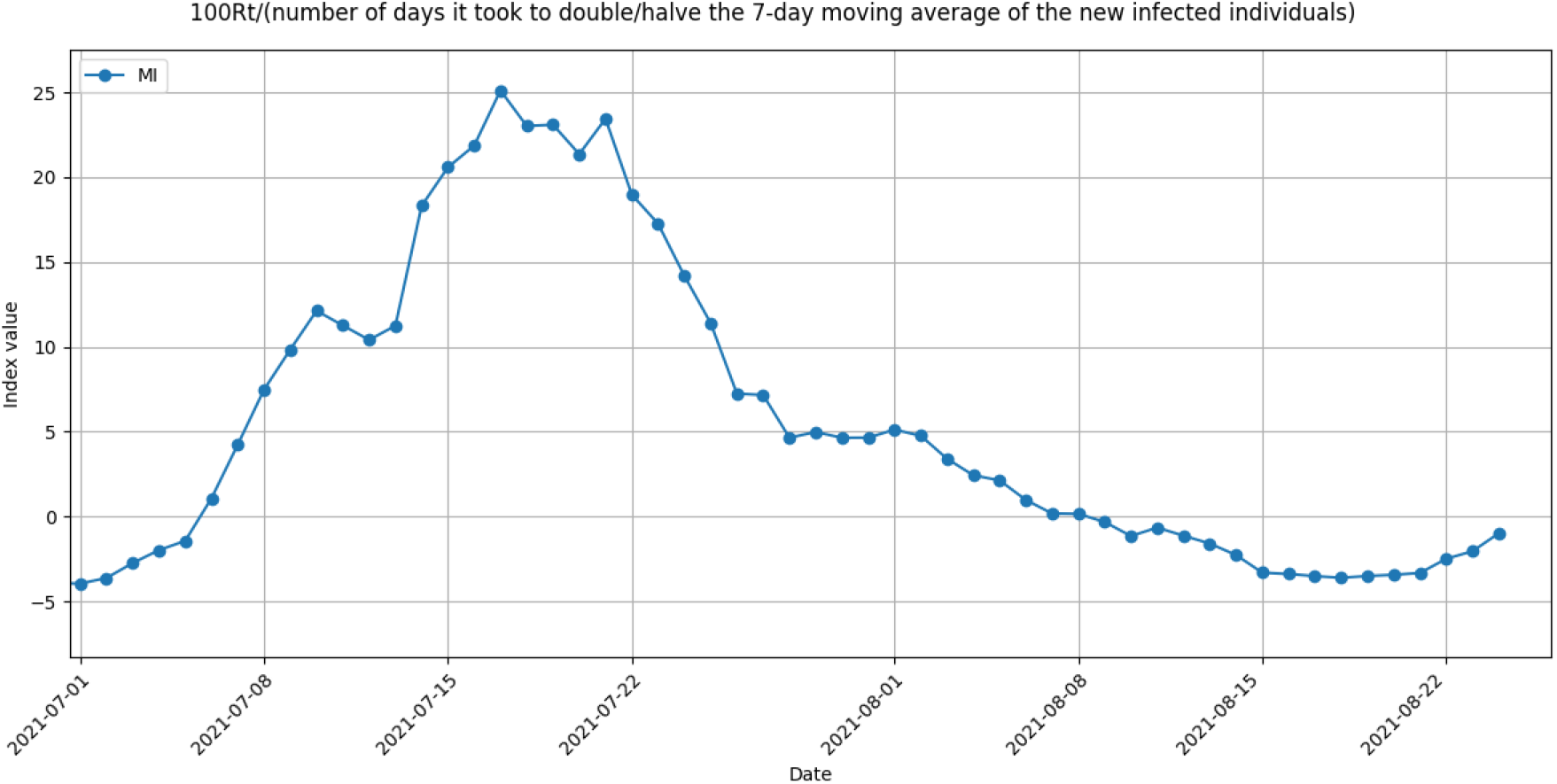
Zoom in of the graph in Fig. 5 for the months of July and August 2021.

This index is practically identical to the RIC-index introduced in [1]. In [1], the authors suggest that abrupt increases of the value of the RIC-index may be precursors of new pandemic waves.

### 3.2 Comparing waves

Figures 7-9 present the graphs of, respectively, the function *R*_*t*_(*I*_*t*_), the number of new infected individuals, the RDT-index for the Italian region of Sicily between August 25th, 2020 and August 24th, 2021. There, four main pandemic waves have occurred in the last year.

**Figure 7:**
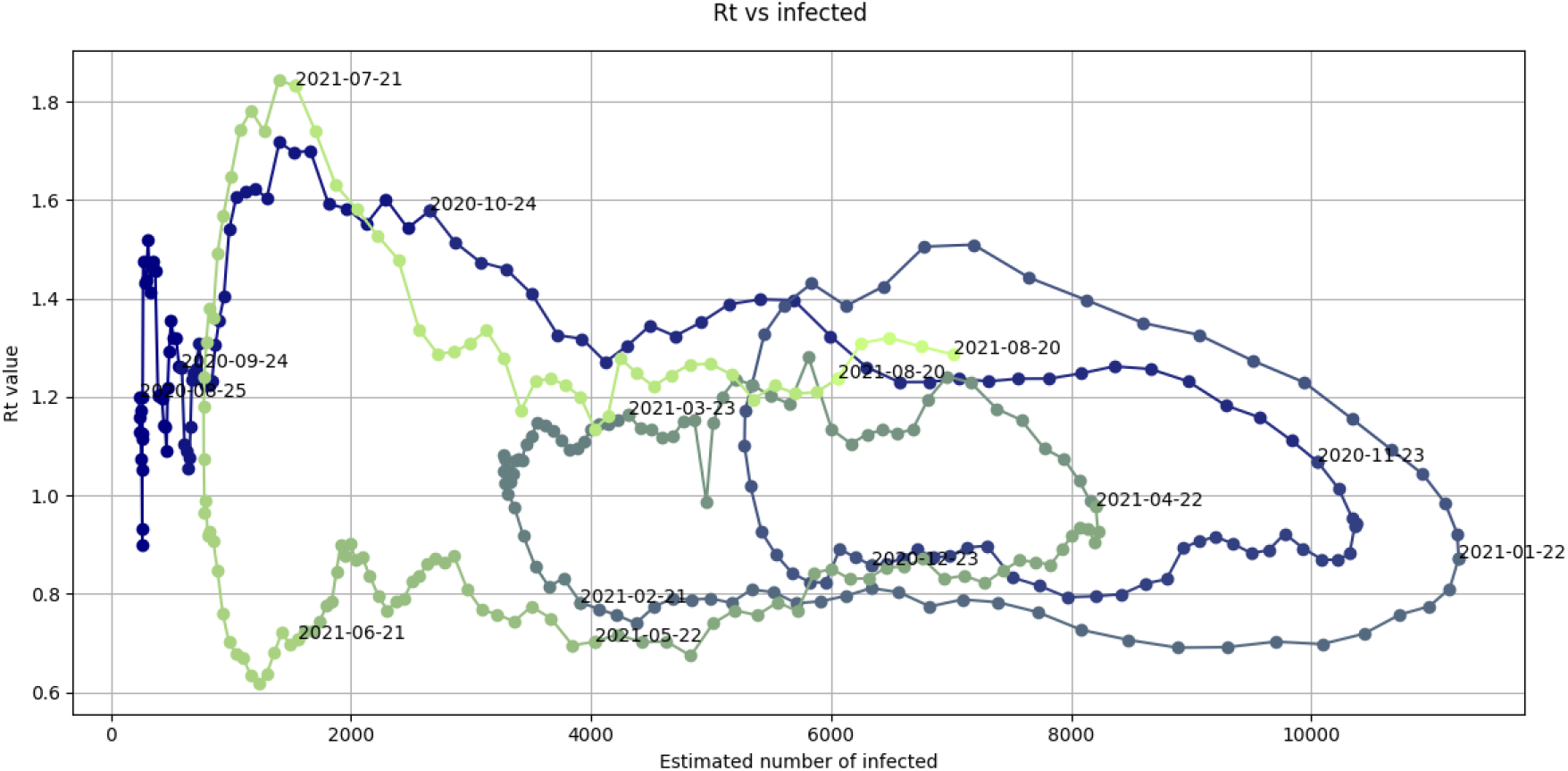
Graph of the values of the *R*_*t*_(*I*_*t*_) function observed in the region of Sicily between August 25th, 2020 and August 24th, 2021.

**Figure 8:**
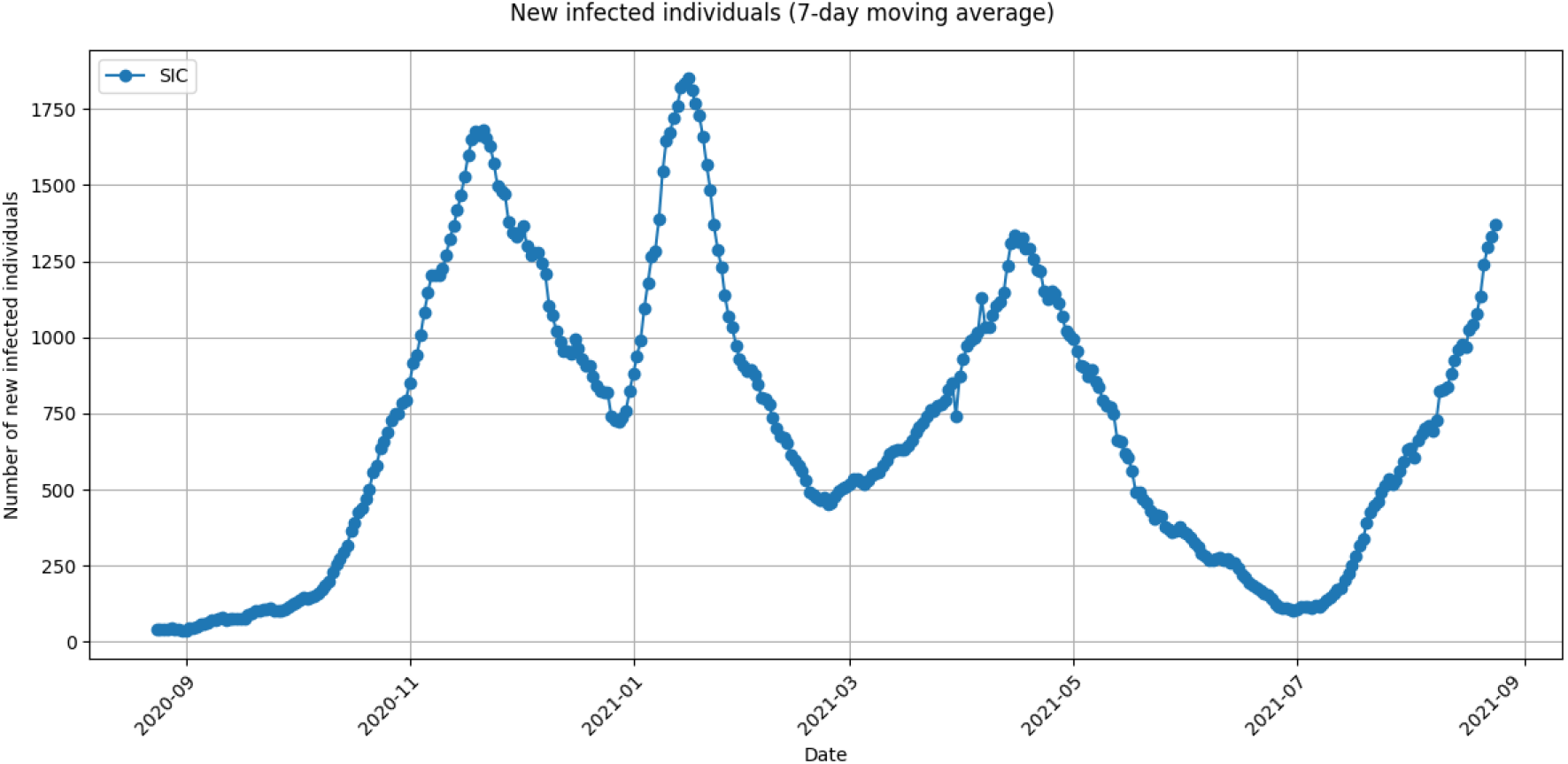
7-day moving average of the number of new infected individuals observed in the region of Sicily between August 25th, 2020 and August 24th, 2021.

**Figure 9:**
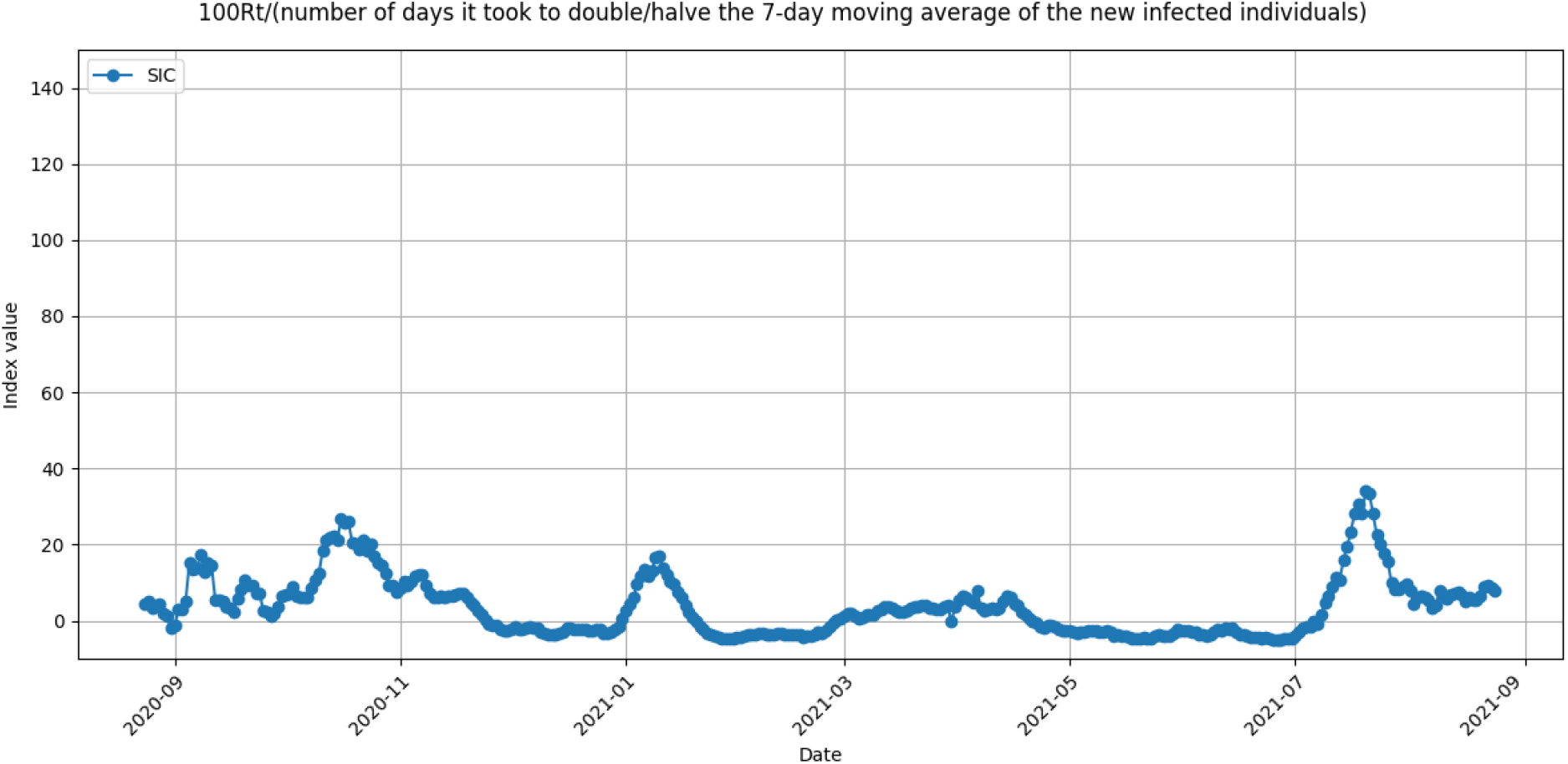
Ratio between *R*_*t*_ and the number of days it took to double/halve the 7-day moving average of the number of new infected individuals observed in the region of Sicily between August 25th, 2020 and August 24th, 2021.

Figure 10, i.e., the zoom in of Fig. 7, allows us to compare the last pandemic wave that started at the very beginning of July, 2021, with the pandemic wave that started in September-October, 2020. Specifically, both waves are preceded by a parallel steep increase of *R*_*t*_, followed by a similar slow decline of this index. We remark that the first measures to limit the October 2020 wave in Sicily wave were adopted on October 24th, 2020. On that day, high-schools were closed, the number of passengers allowed to board trains and buses were reduced by 50% and a night curfew was imposed. Differently, no measure is imposed to the date of August 25th, 2021, to limit the effects of the last wave.

**Figure 10:**
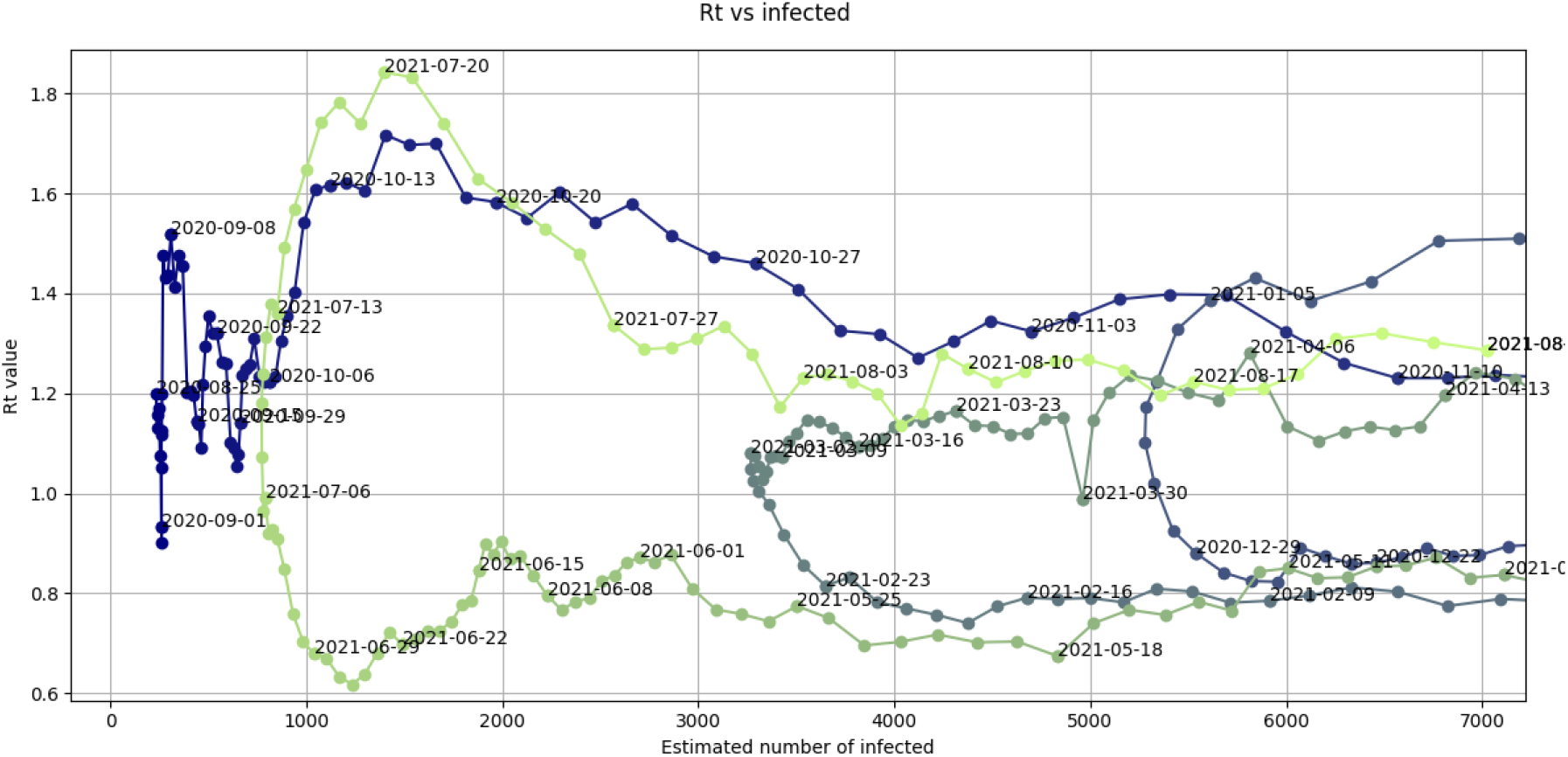
Zoom in of the graph in Fig. 7 where the values of the *R*_*t*_(*I*_*t*_) function in the months of July and August 2021 are highlighted.

### 3.3 Phases

If we accept a certain level of arbitrariness, we can distinguish four phases, plus possibly an initial one (hereinafter referred as to Phase 0), in the loops of the graph of function *R*_*t*_(*I*_*t*_).

#### 3.3.1 Phase 0

Phase 0 is characterized by strong oscillations, if not one or more outbursts, of the *R*_*t*_ value in presence of small number of infected individuals, (see, e.g., Fig. 11 and its zoom in Fig. 12).

**Figure 11:**
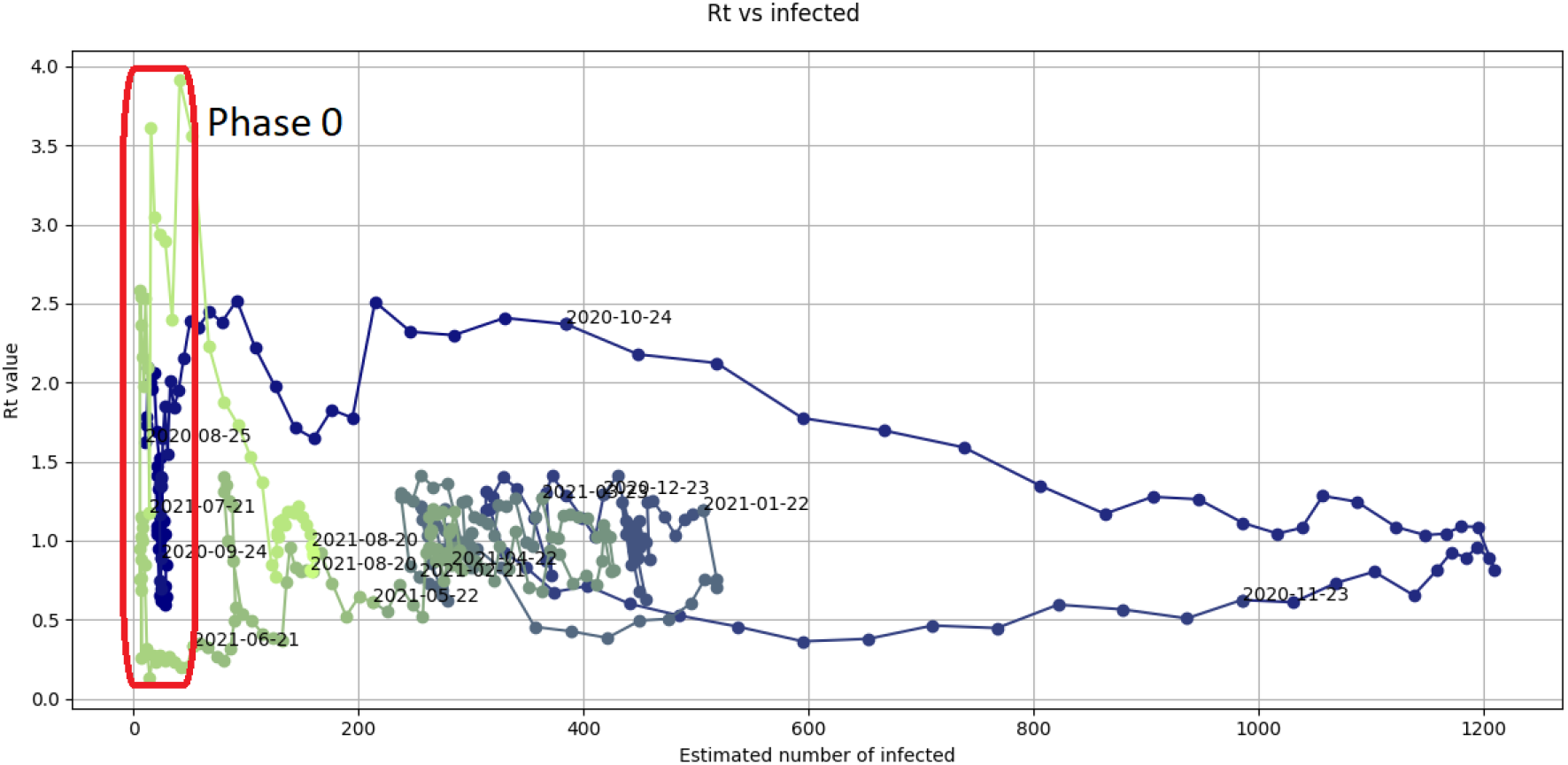
Graph of the values of the *R*_*t*_(*I*_*t*_) function observed in the province of Viterbo between August 25th, 2020 and August 24th, 2021. Phases 0 are highlighted.

**Figure 12:**
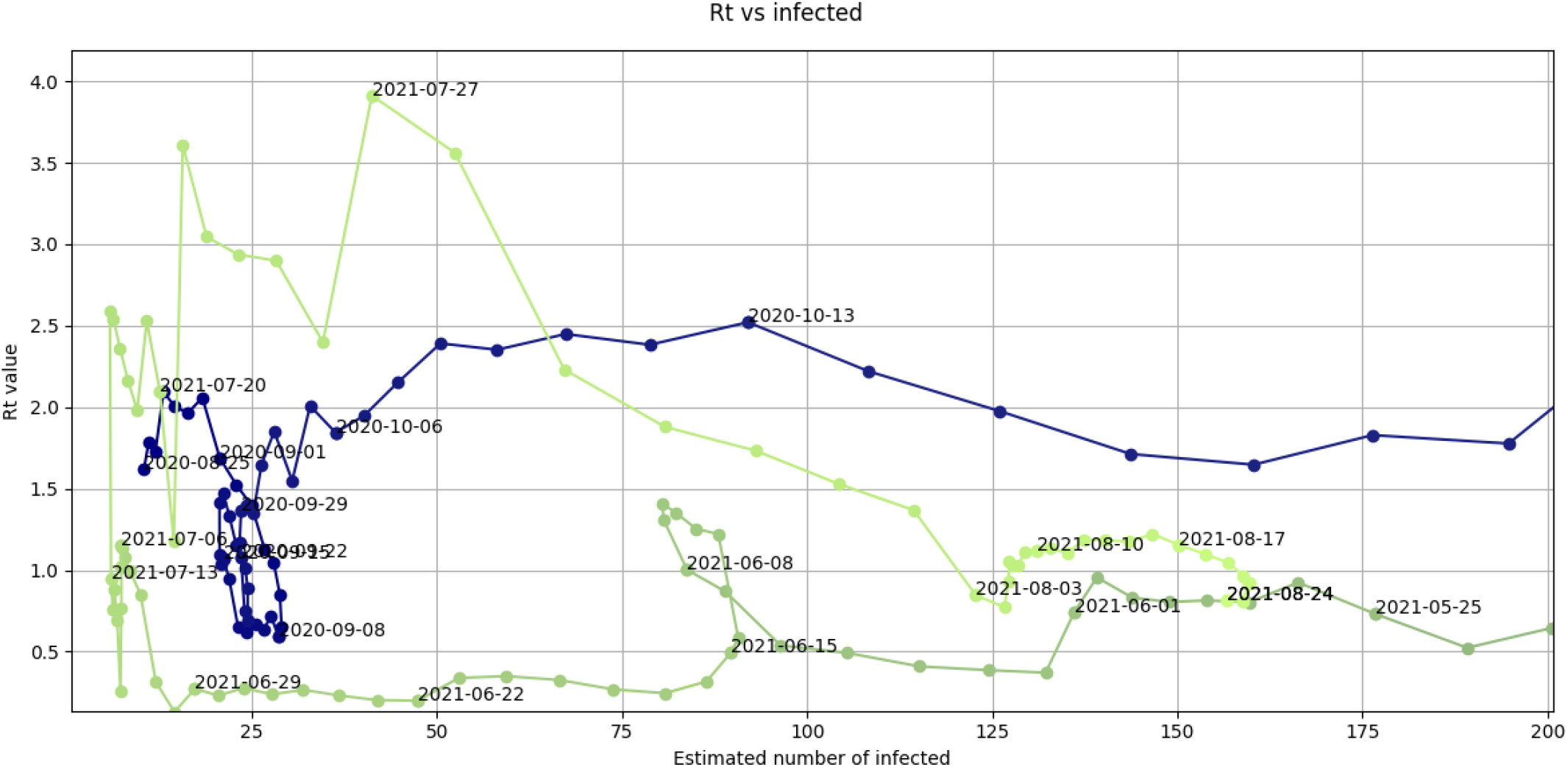
Zoom in of the graph in Fig. 11 for low numbers of estimated infected individuals.

Two main reasons justify the above pattern:

- occurrence of some outbreaks, where few infected individuals caused many new infected individuals possibly due to their participation to, e.g., some social events;
- errors in data collection or recording.

Periods of strong oscillations in the RIC-index graph (see, e.g., Fig. 13) correspond to Phases 0 in the graph of function *R*_*t*_(*I*_*t*_). The periods of occurrence of these oscillations in the two graphs may coincide (see, e.g., Fig. 14) or Phases 0 may appear slightly earlier (see, e.g., Fig. 15).

**Figure 13:**
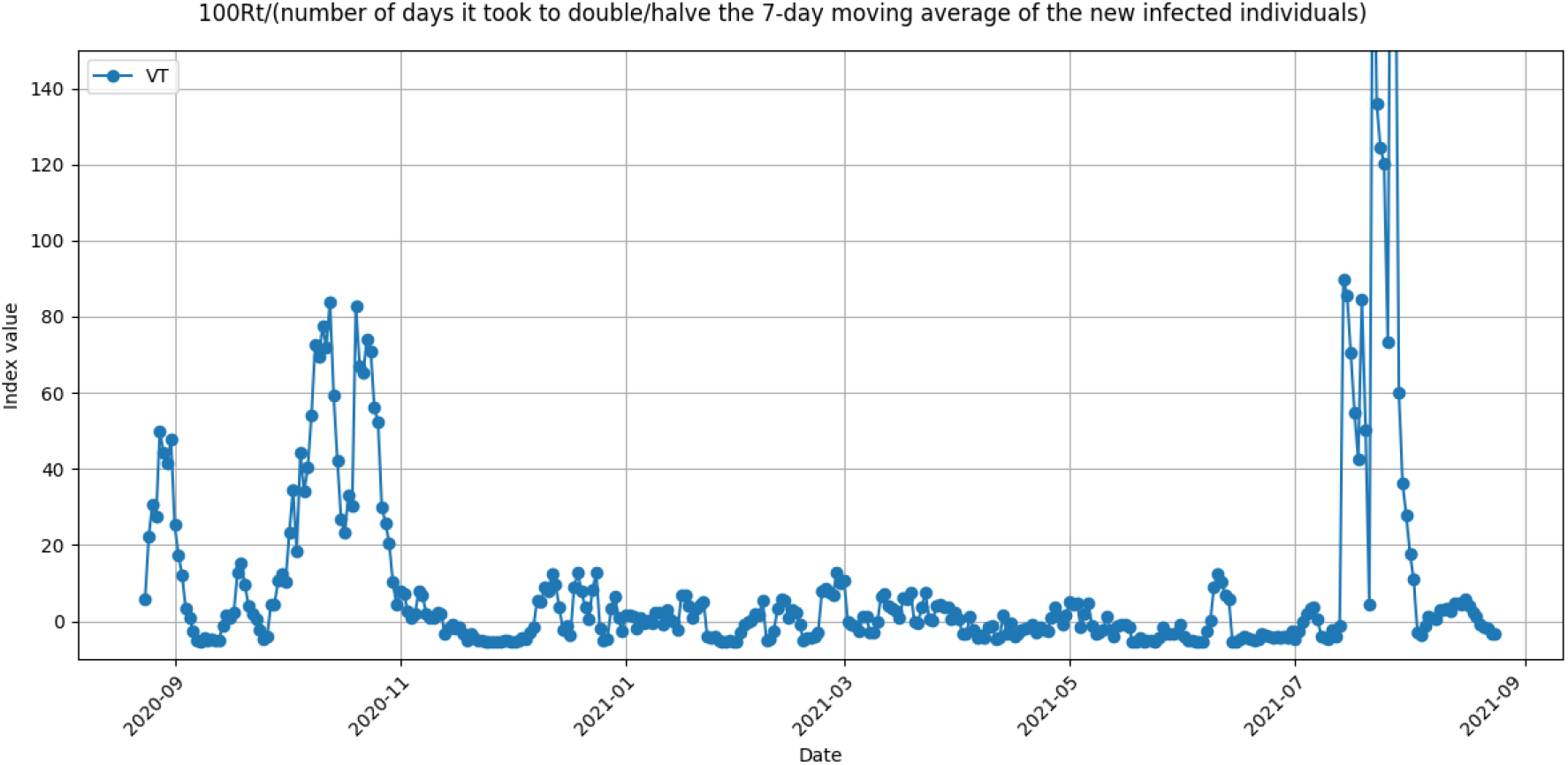
Ratio between *R*_*t*_ and the number of days it took to double/halve the 7-day moving average of the number of new infected individuals observed in the province of Viterbo between August 25th, 2020 and August 24th, 2021.

**Figure 14:**
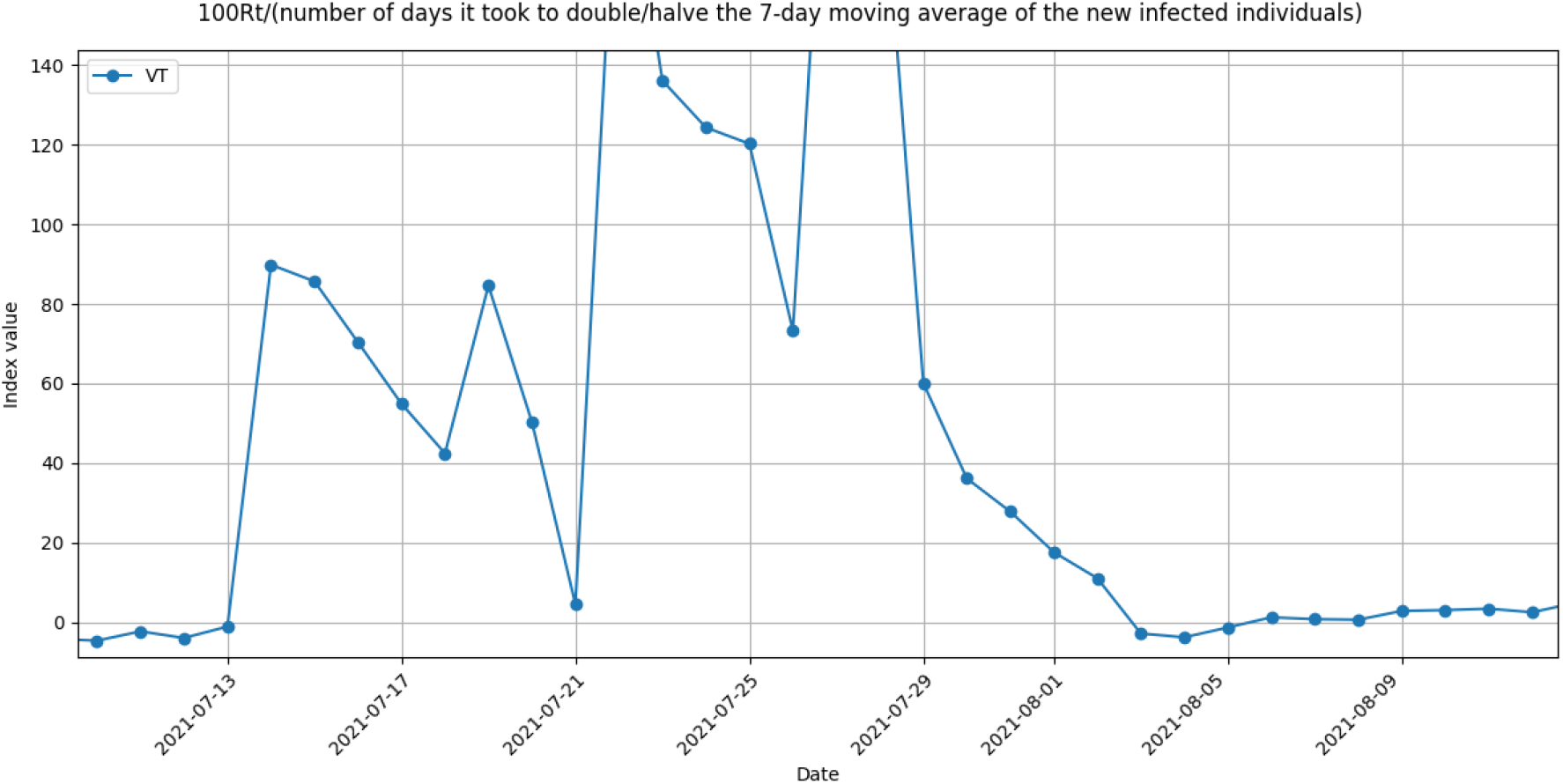
Zoom in of the graph in Fig. 13 for the months July and August, 2021.

**Figure 15:**
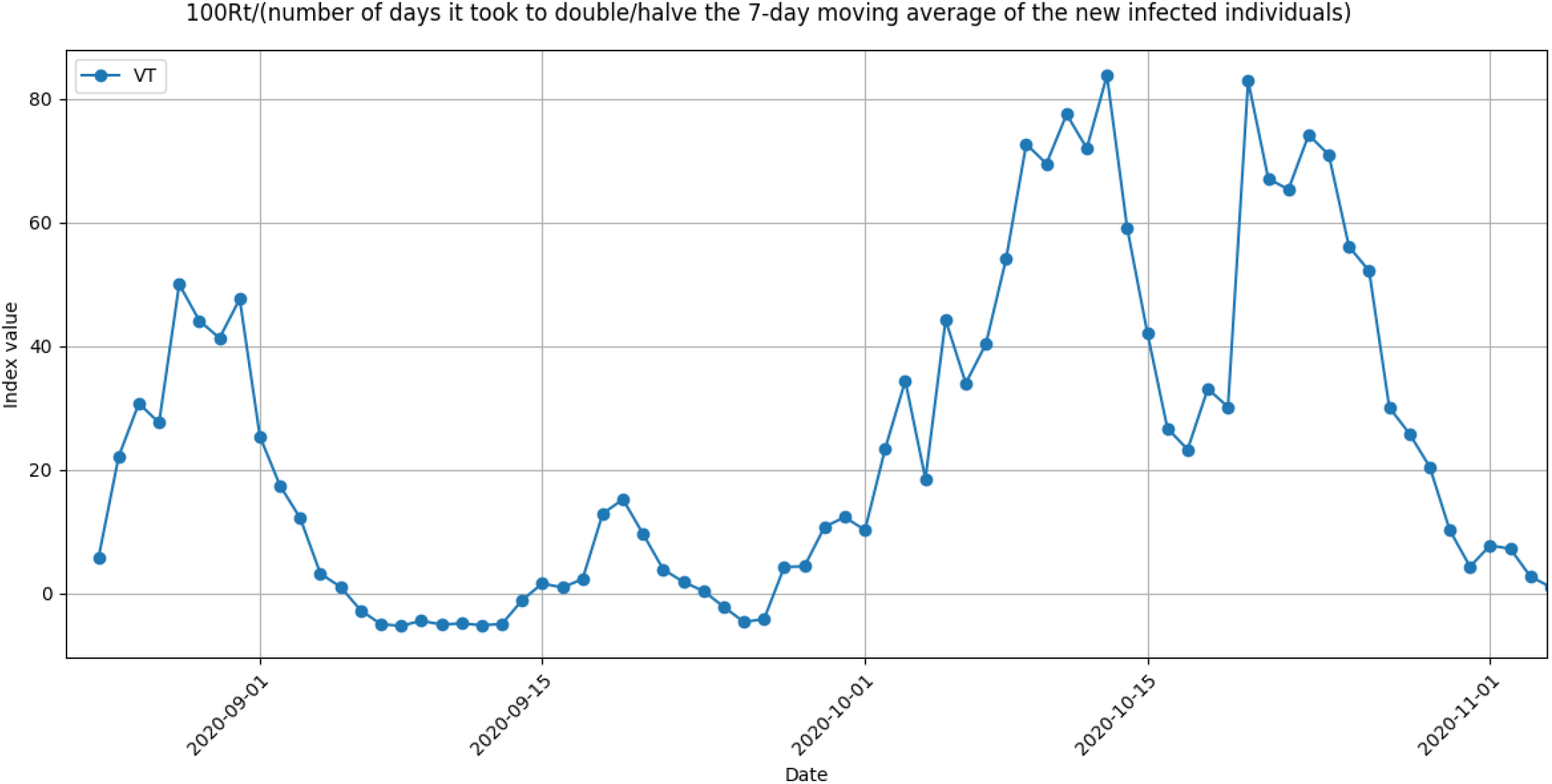
Zoom in of the graph in Fig. 13 for the months September and October, 2020.

Needless to say that the occurrence of a Phase 0 does not necessarily imply a new pandemic wave if immediate measures to isolate the infected individuals involved in the outbreaks are implemented. Apparently, one can deduce from Fig. 12 and Fig. 16 that authorities were not able to contain the relatively small outbreaks in September and October, 2020, whereas so far they succeeded in July and August, 2021.

**Figure 16:**
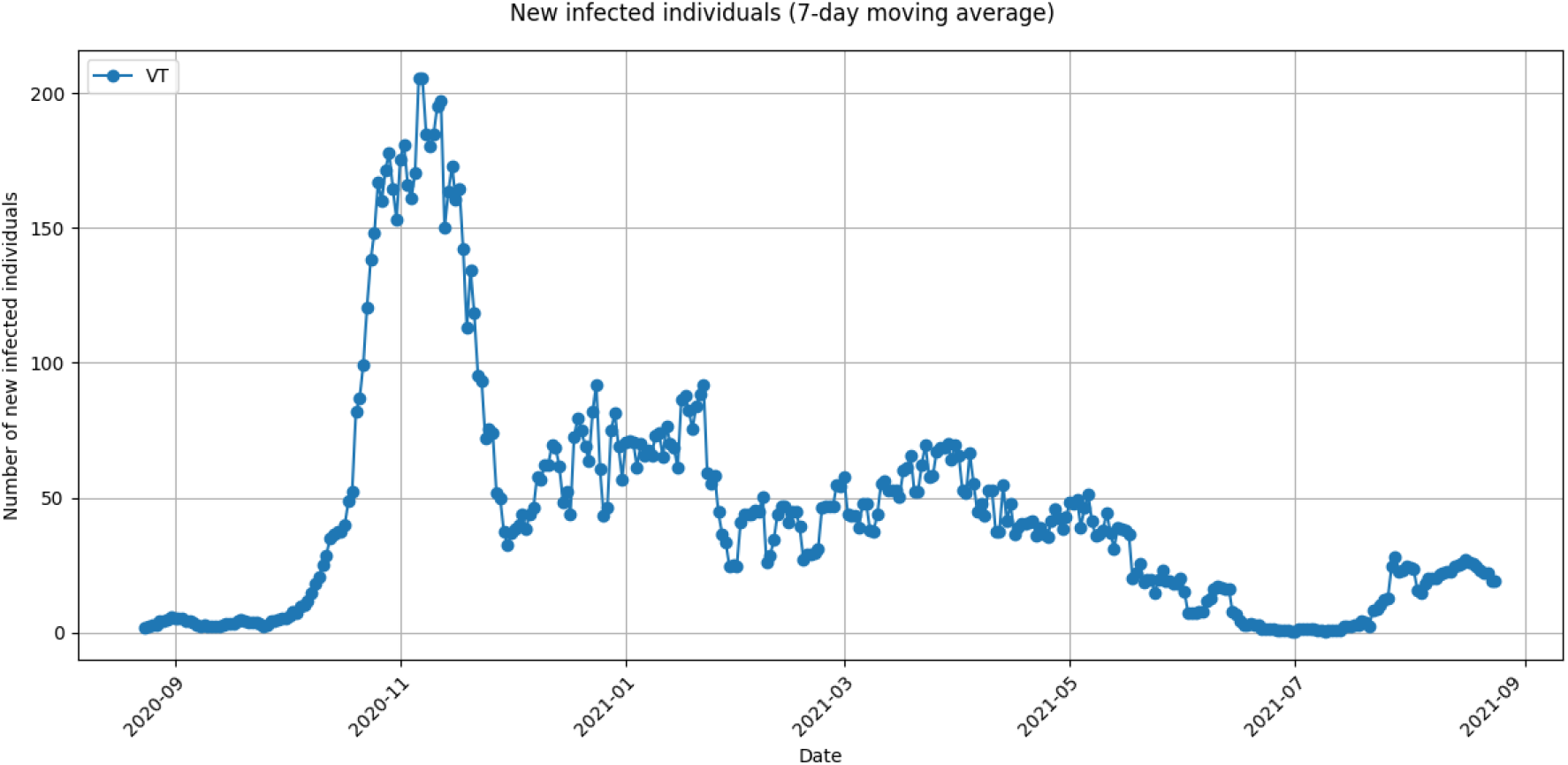
7-day moving average of the number of new infected individuals observed in the province of Viterbo between August 25th, 2020 and August 24th, 2021.

Figures 17-20 presents graphs of function *R*_*t*_(*I*_*t*_) with an evident Phase 0 for Italian provinces of characterized by different amount of population:

**Figure 17:**
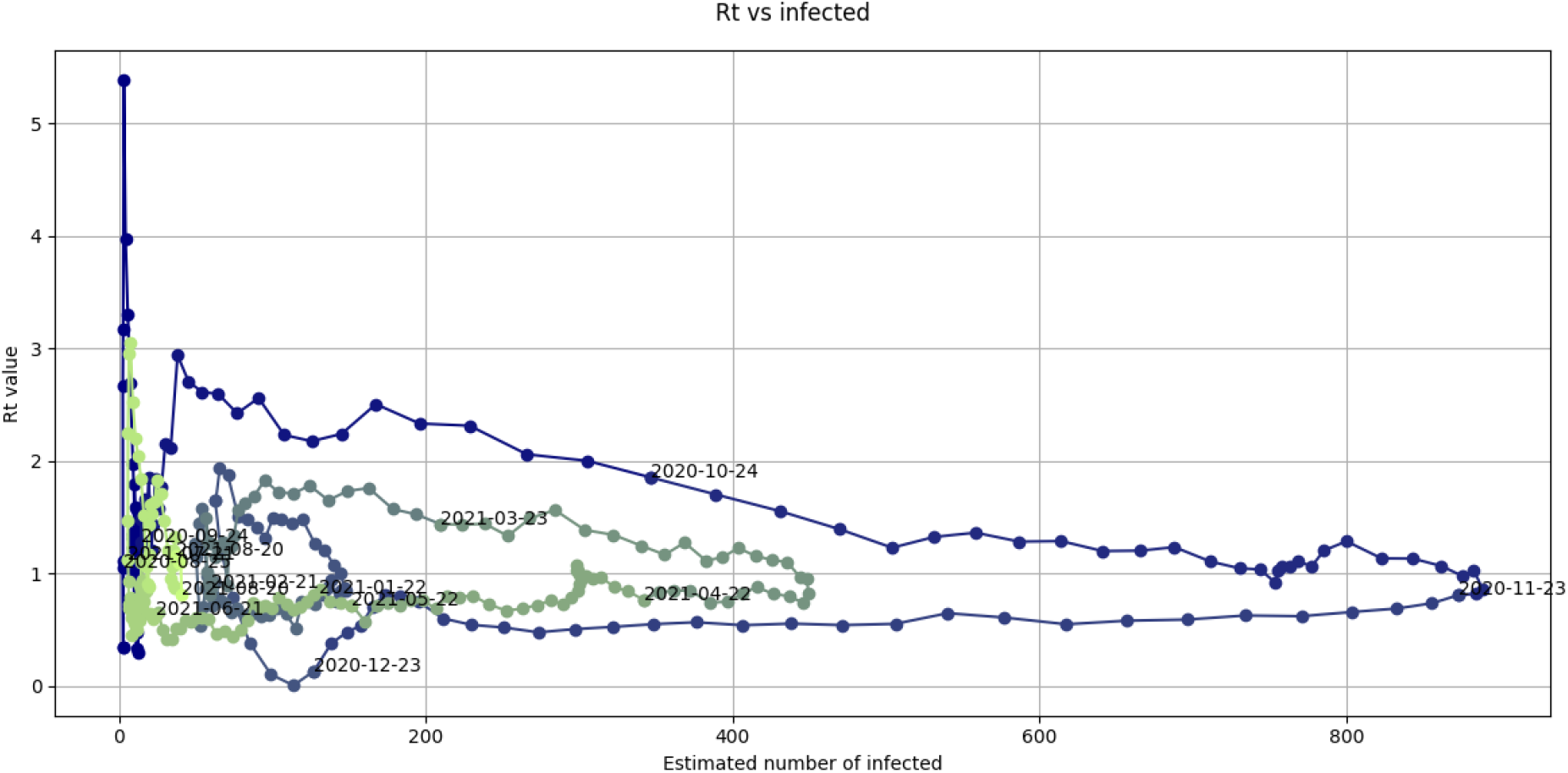
Graph of the values of the *R*_*t*_(*I*_*t*_) function observed in the province of Aosta between August 25th, 2020 and August 24th, 2021.

**Figure 18:**
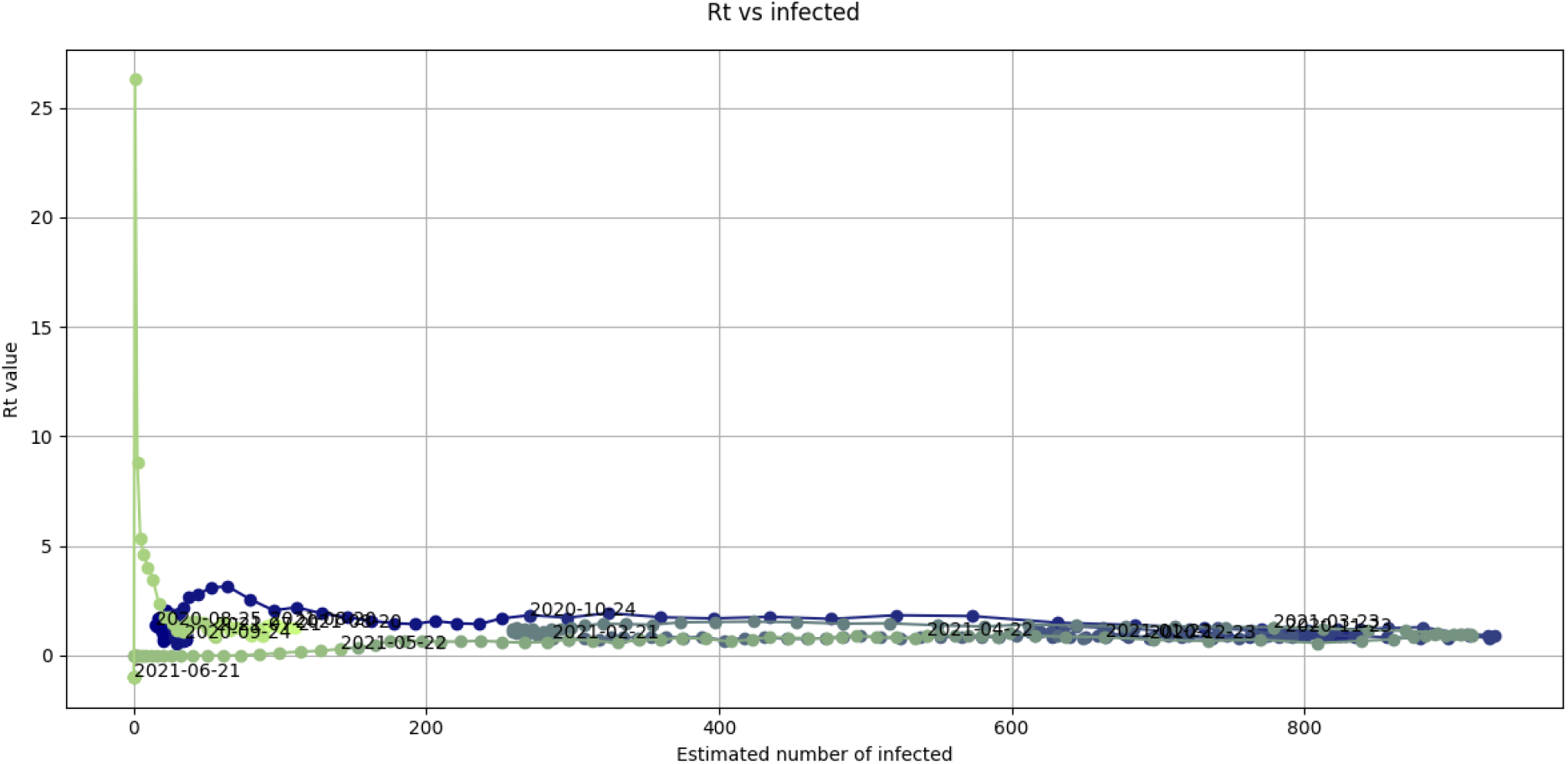
Graph of the values of the *R*_*t*_(*I*_*t*_) function observed in the province of Trieste between August 25th, 2020 and August 24th, 2021.

**Figure 19:**
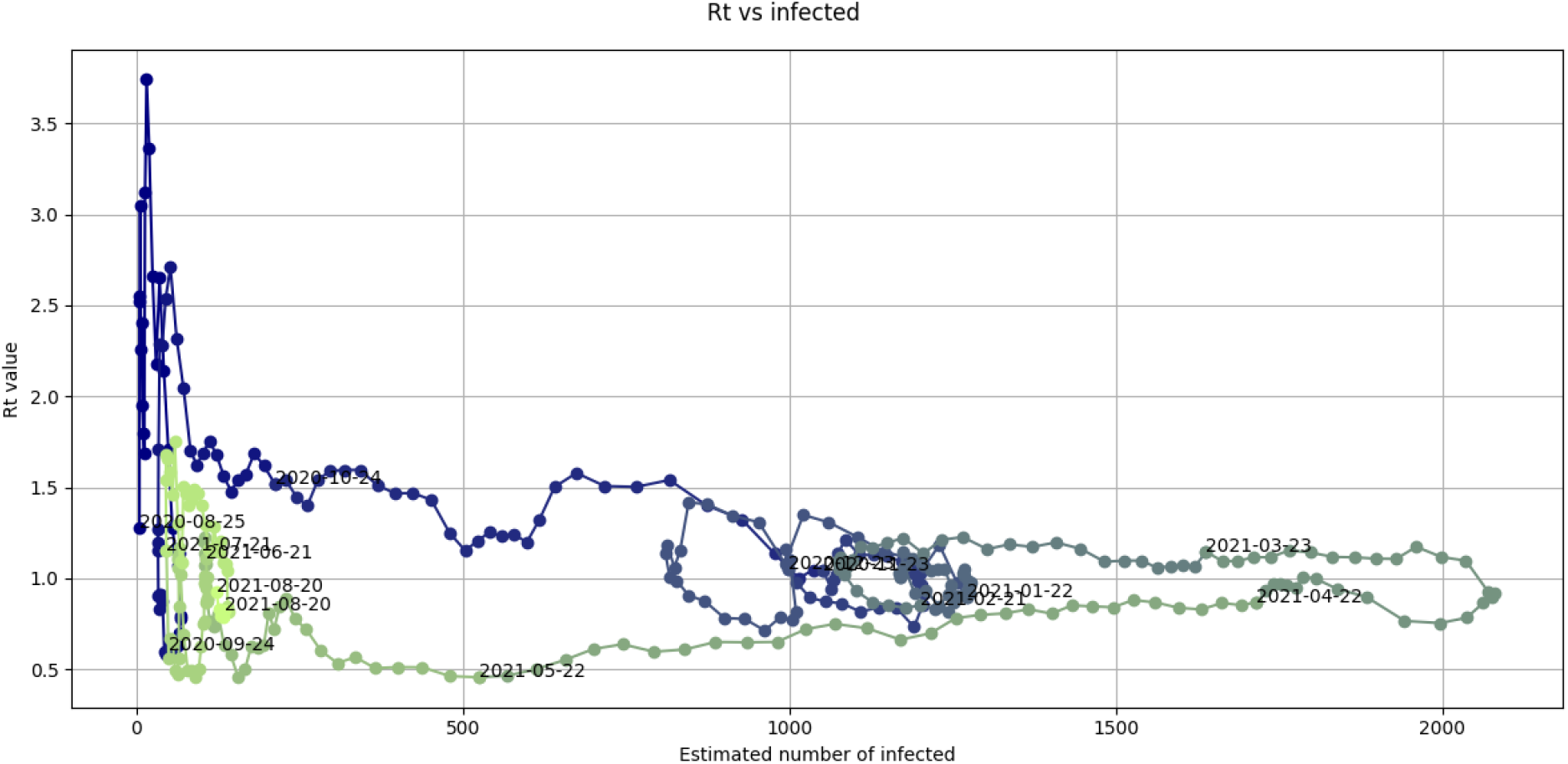
Graph of the values of the *R*_*t*_(*I*_*t*_) function observed in the province of Taranto between August 25th, 2020 and August 24th, 2021.

**Figure 20:**
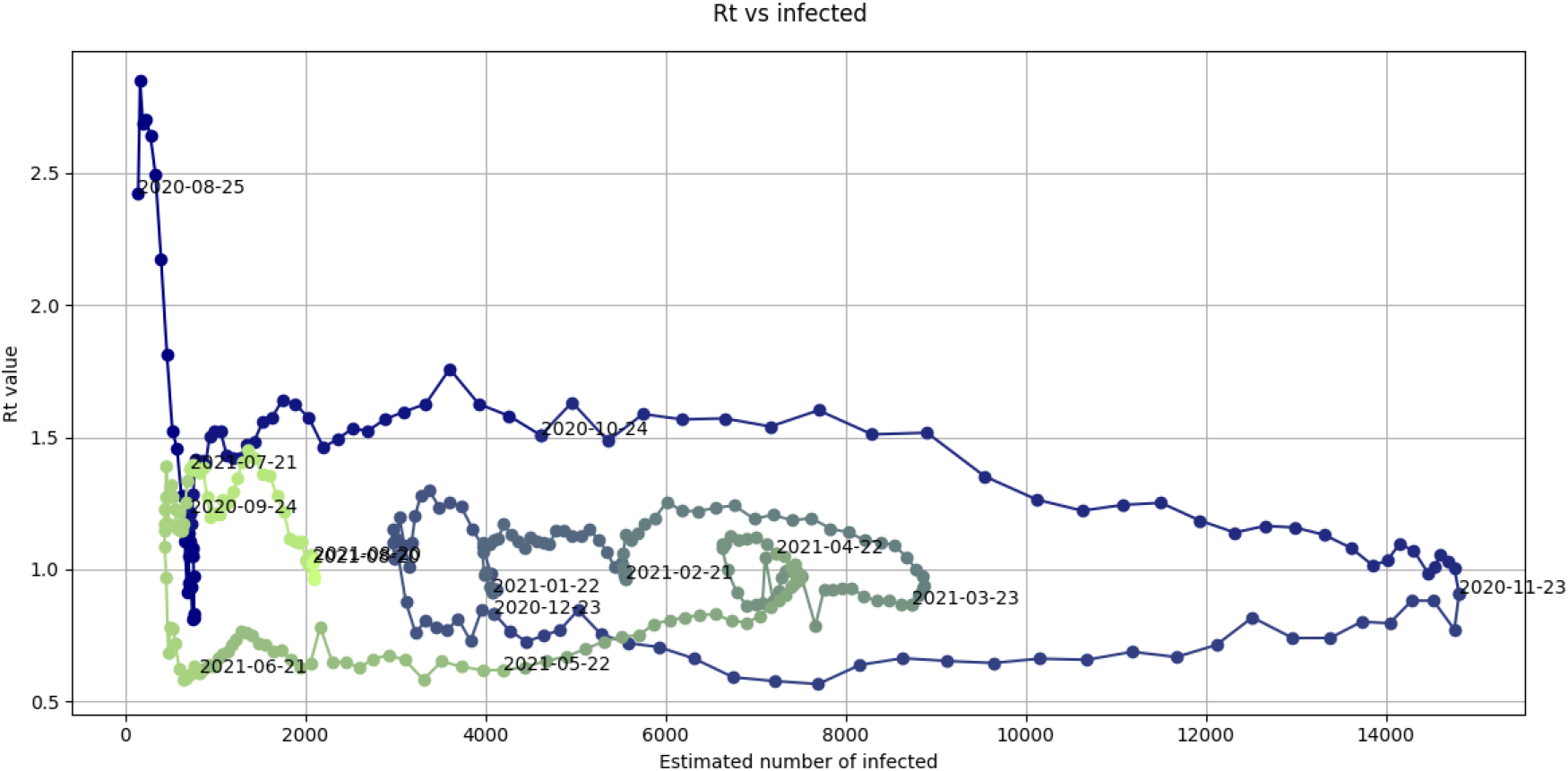
Graph of the values of the *R*_*t*_(*I*_*t*_) function observed in the province of Naples between August 25th, 2020 and August 24th, 2021.

- Aosta: about 124.000 inhabitants (the second least populated province of Italy)
- Trieste: about 230.000 inhabitants
- Taranto: about 560.000 inhabitants
- Napoli: about 3.017.000 inhabitants (the third most populated province of Italy)

#### 3.3.2 Phase 1

Phase 1 is characterized by an horizontal trend or a slight decline of the *R*_*t*_ value in presence of an increasing number of infected individuals (see Fig. 21 and Fig. 22), with a possible stabilization of *R*_*t*_ to a value greater than 1.

**Figure 21:**
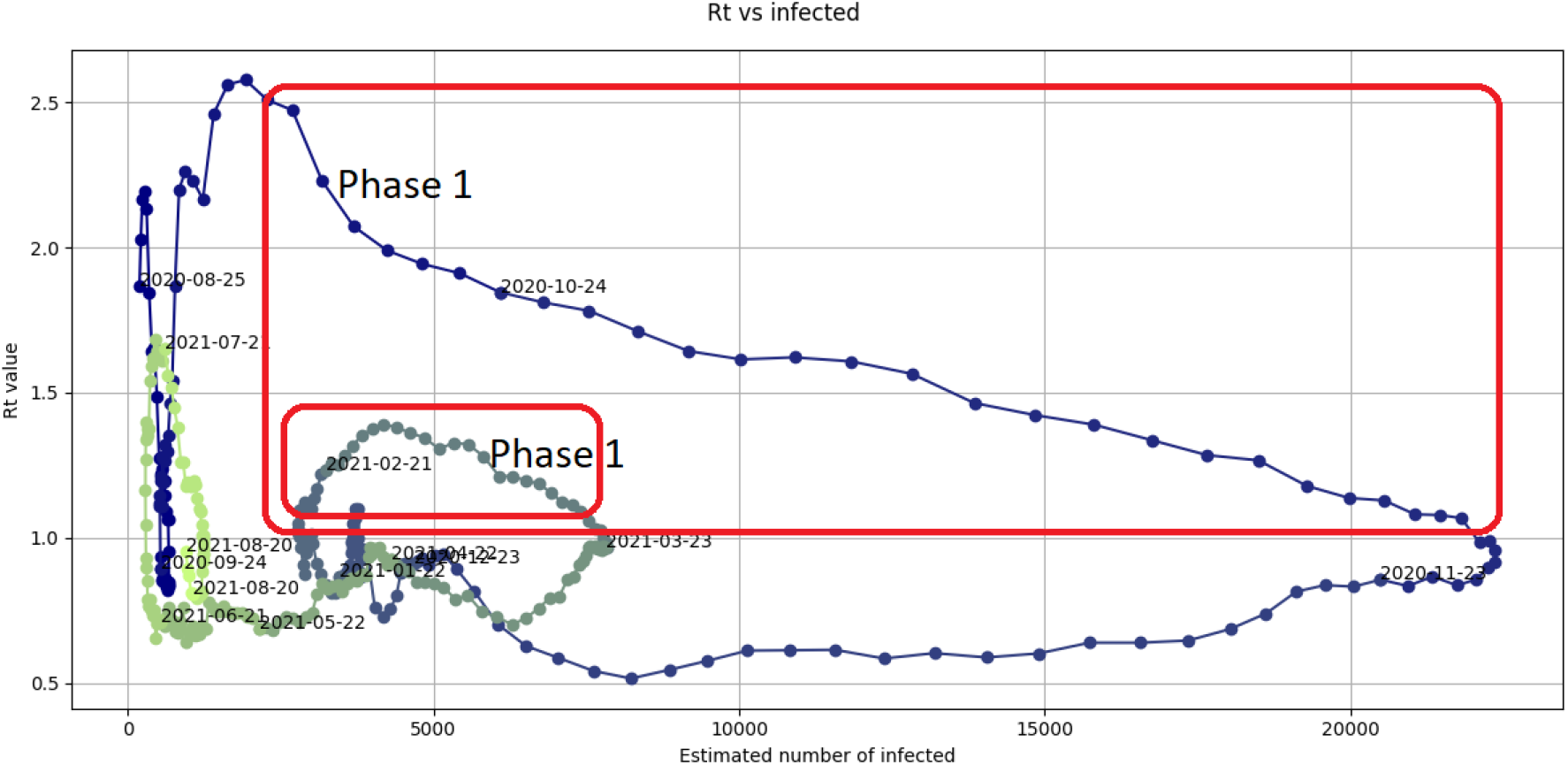
Graph of the values of the *R*_*t*_(*I*_*t*_) function observed in the province of Milan between August 25th, 2020 and August 24th, 2021. Phases 1 are highlighted.

**Figure 22:**
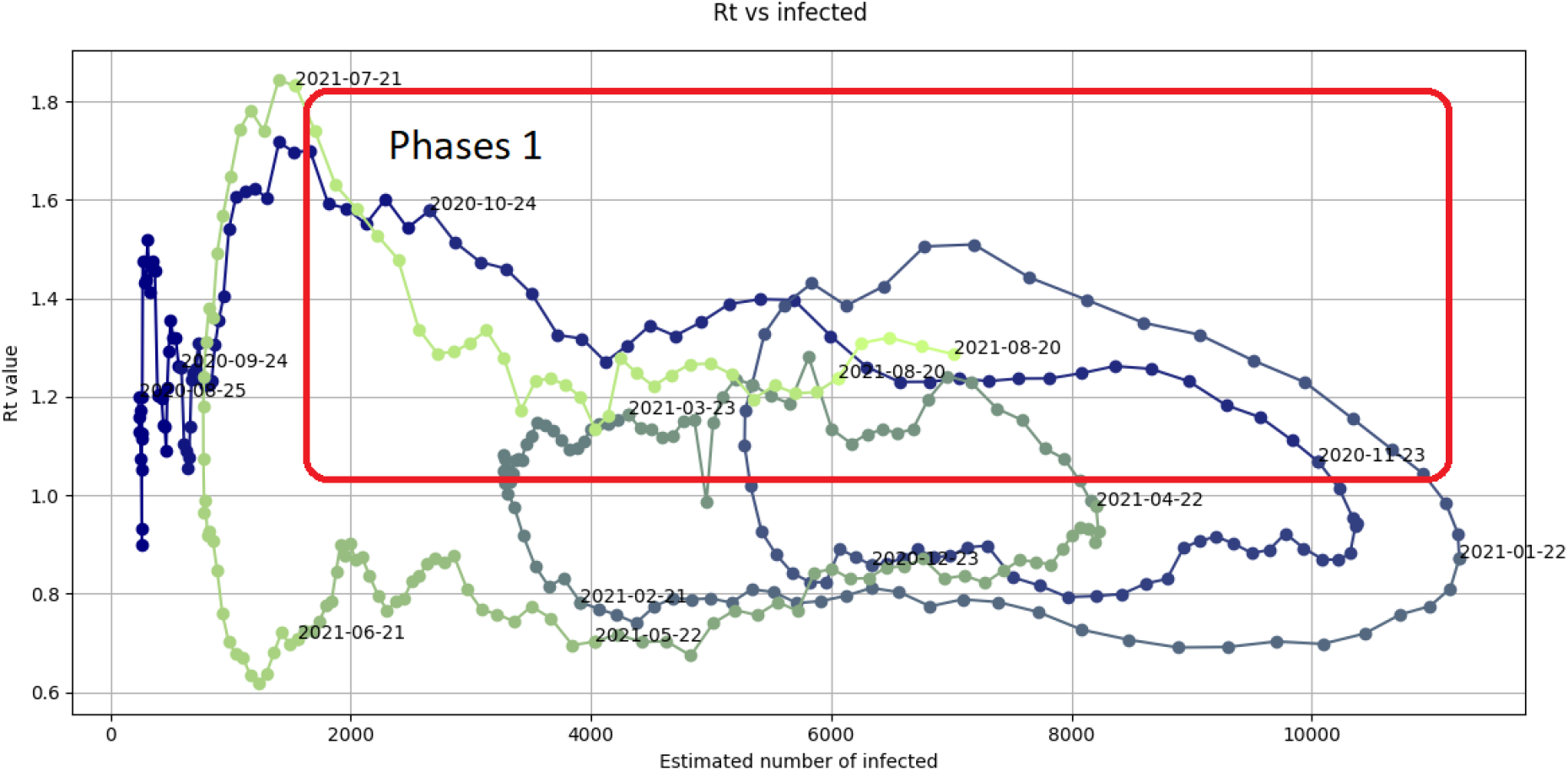
Graph of the values of the *R*_*t*_(*I*_*t*_) function observed in the region of Sicily between August 25th, 2020 and August 24th, 2021. Phases 1 are highlighted.

A Phase 1 corresponds to the growth of a pandemic wave in the new infected individuals graph. The longer (along the abscissa axis) is a Phase 1, the higher is a pandemic wave. The higher is the average value of *R*_*t*_ in a Phase 1, the steeper is a pandemic wave.

Both Fig. 21 and Fig. 22 show a relative rapid decrease of the *R*_*t*_ value during Phases 1 if the *R*_*t*_ has reached a high value. As an example, the province of Milan adopted the first containment measures for the October 2020 wave on October 22nd, when an initial decrease of the *R*_*t*_ value had already occurred (see Fig. 21). The region of Sicily adopted almost no special measures to contrast the July-August 2021 wave, which indeed is greater than in other Italian regions, but the *R*_*t*_ value decreased all the same. Possibly, the population spontaneously adopts a precautionary behavior when a high intensity of risk is perceived. Possibly, a good communication strategy by the authorities may foster this behavior.

#### 3.3.3 Phase 2

Phase 2 is characterized by a steep, some times almost vertical, drop of the *R*_*t*_ value that goes below 1 in presence of a relative stability in the number of infected individuals (see Fig. 23-25),

**Figure 23:**
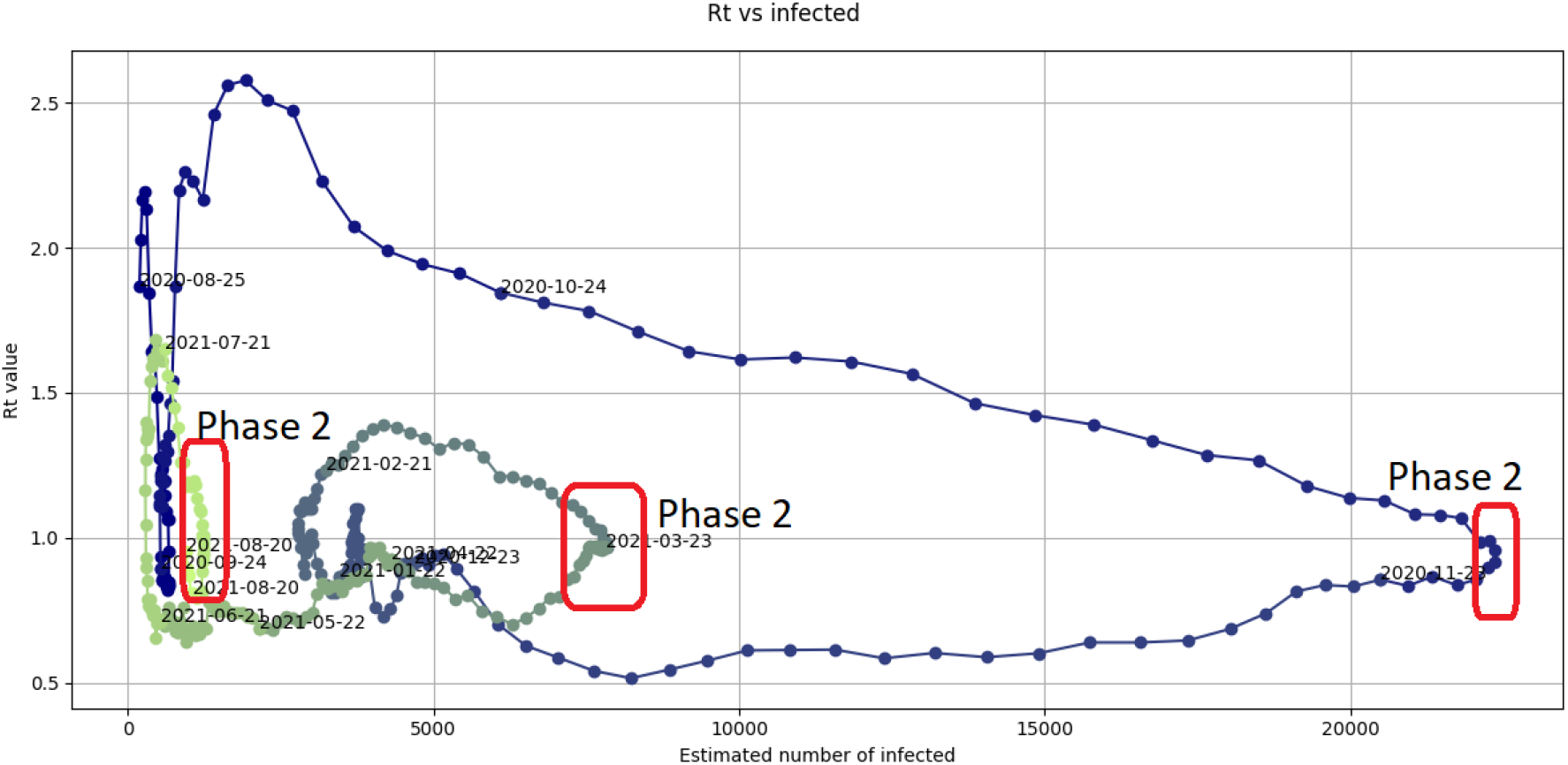
Graph of the values of the *R*_*t*_(*I*_*t*_) function observed in the province of Milan between August 25th, 2020 and August 24th, 2021. Phases 2 are highlighted.

These phases may be associated to:

- seasonal trend of the pandemic;
- results of the implementation of containment measures;
- special local characteristics.

Consequently, these phases may or may not occur at the same time in the different Italian regions and provinces.

As an example, we can observe a Phase 2 around November 22nd, 2020, in the graphs of all three Fig. 23-25. Differently, only the province of Rome (the largest Italian province) and Milan present a Phase 2 in August, 2021, (see Fig. 23 and Fig. 24) possibly, because the percentage of vaccinated is less in Sicily (see Fig. 25) than in the provinces of Rome and Milan, or because Sicily is a holiday destination in August.

**Figure 24:**
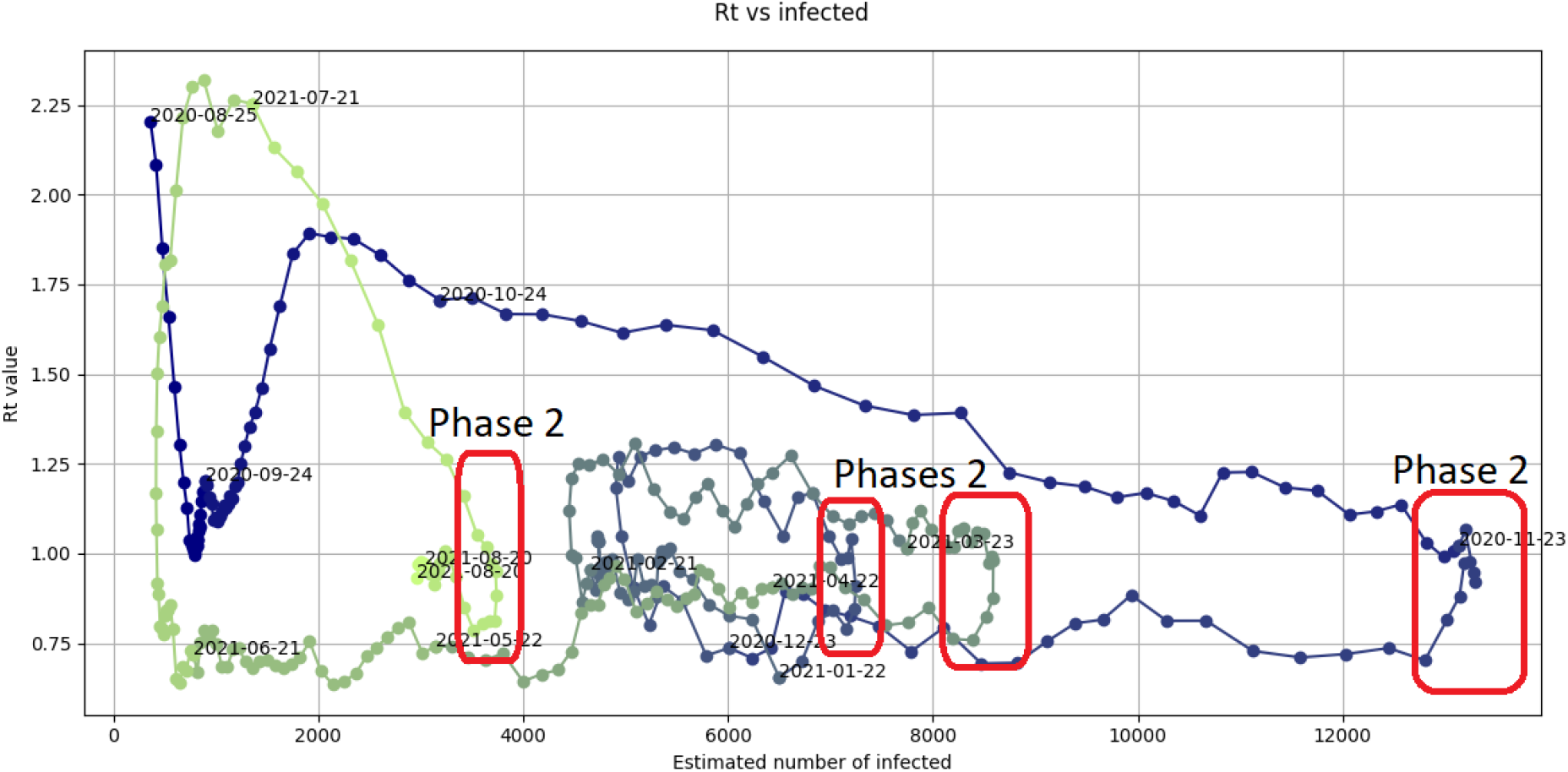
Graph of the values of the *R*_*t*_(*I*_*t*_) function observed in the province of Rome between August 25th, 2020 and August 24th, 2021. Phases 2 are highlighted.

**Figure 25:**
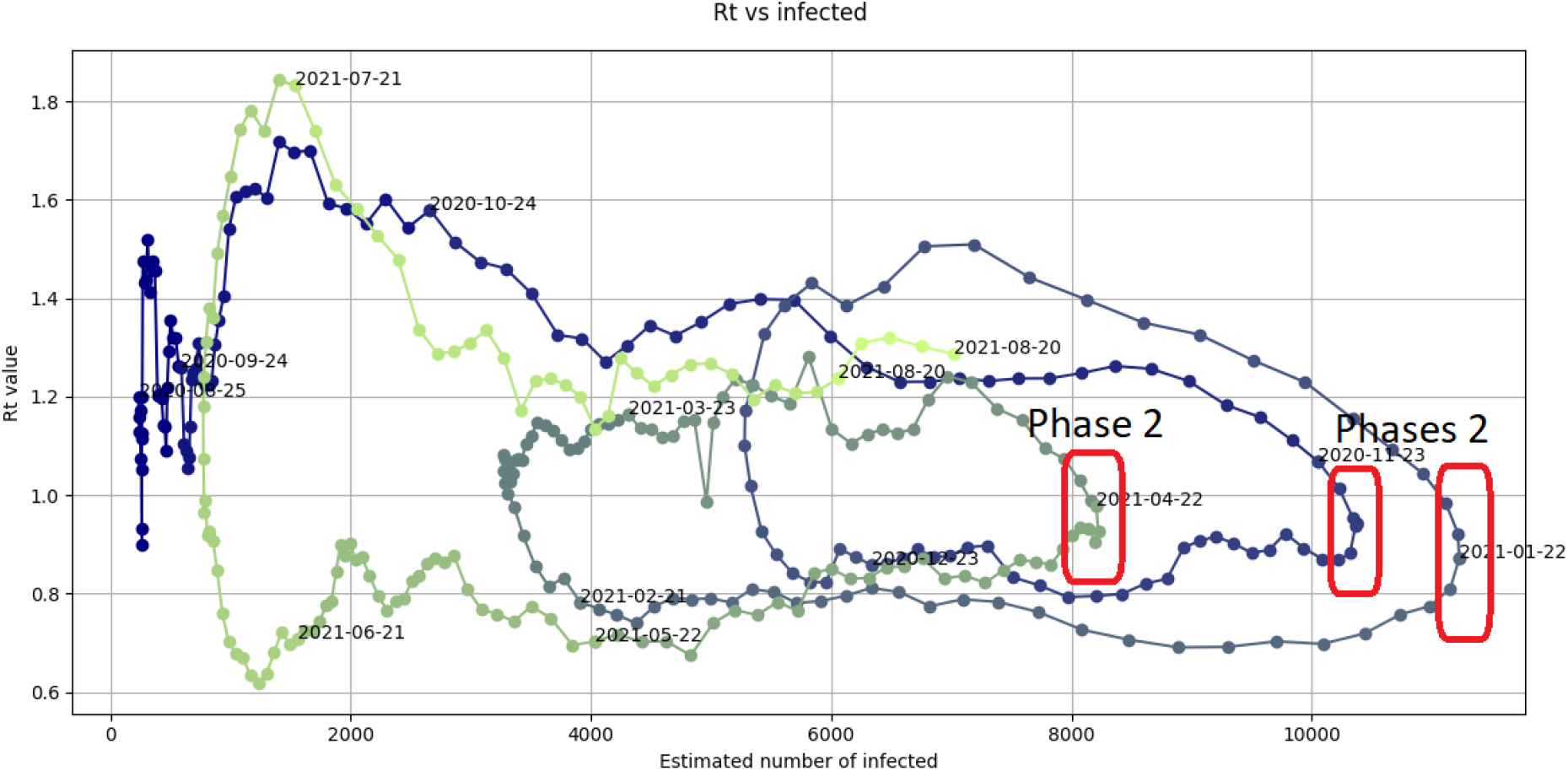
Graph of the values of the *R*_*t*_(*I*_*t*_) function observed in the region of Sicily between August 25th, 2020 and August 24th, 2021. Phases 2 are highlighted.

**Figure 26:**
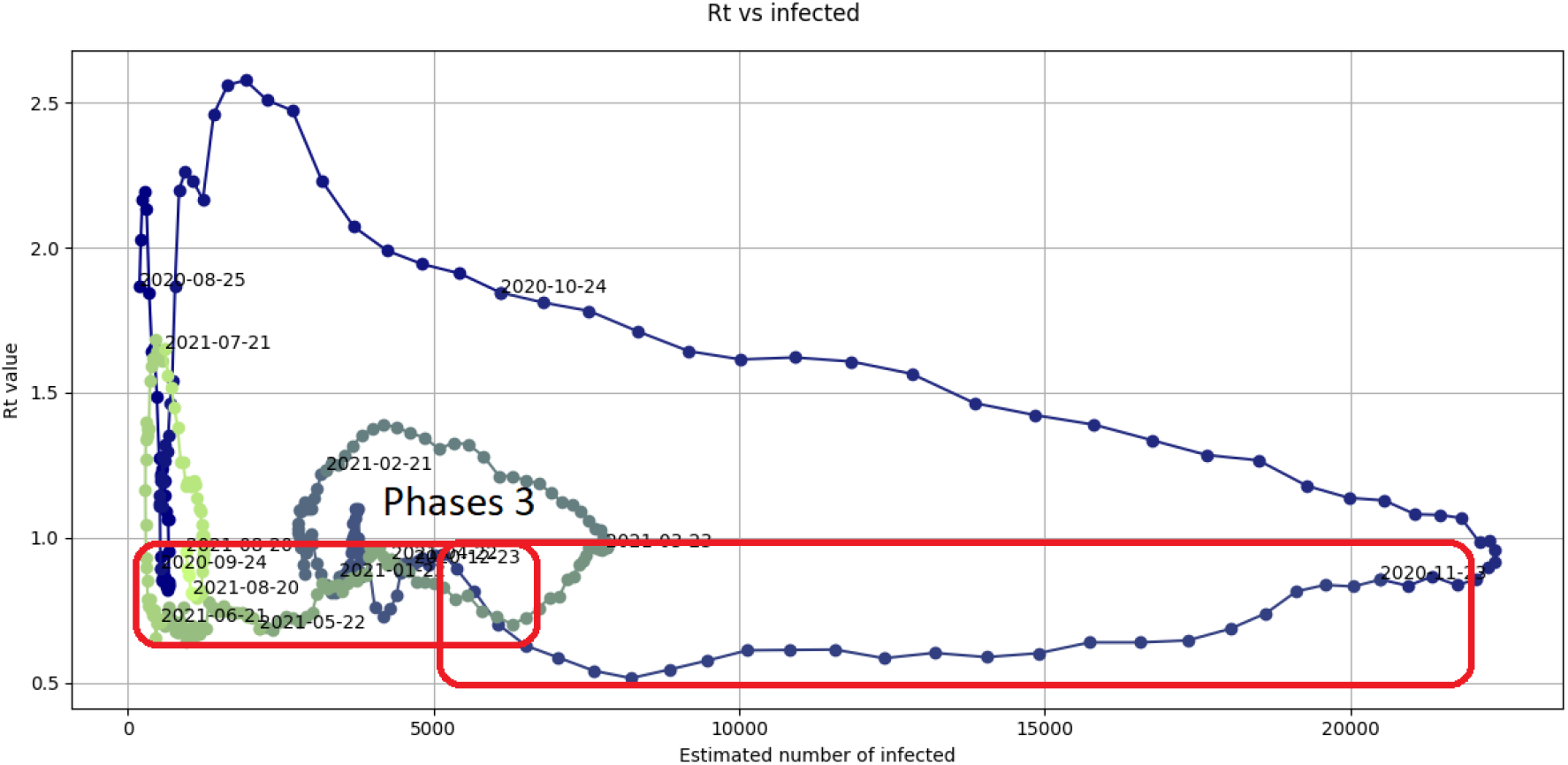
Graph of the values of the *R*_*t*_(*I*_*t*_) function observed in the province of Milan between August 25th, 2020 and August 24th, 2021. Phases 3 are highlighted.

A Phase 2 corresponds to the period that goes from the slowdown of a pandemic wave growth to its initial decline in the new infected individuals graph.

#### 3.3.4 Phase 3

Phase 3 is characterized by an almost constant *R*_*t*_ value below 1 corresponding to a decrease in the number of new infected individuals.

A Phase 3 corresponds to the decline of a pandemic wave in the new infected individuals graph. The longer (along the abscissa axis) is a Phase 3, the lower level of a pandemic wave is reached. The lower is the average value of *R*_*t*_ in a Phase 3, the steeper is the decline of a pandemic wave.

Theoretically, a Phase 3 could lead to the conclusion of the COVID-19 pandemic. Unfortunately, in Italy, Phases 3 had always followed by either a Phase 0 or 4 so far, i.e., by a new pandemic wave.

#### 3.3.5 Phase 4

Phase 4 is characterized by a steep, some times almost vertical, increase of the *R*_*t*_ value that goes above 1 in presence of a relative stability in the number of infected individuals (see Fig. 27-29).

**Figure 27:**
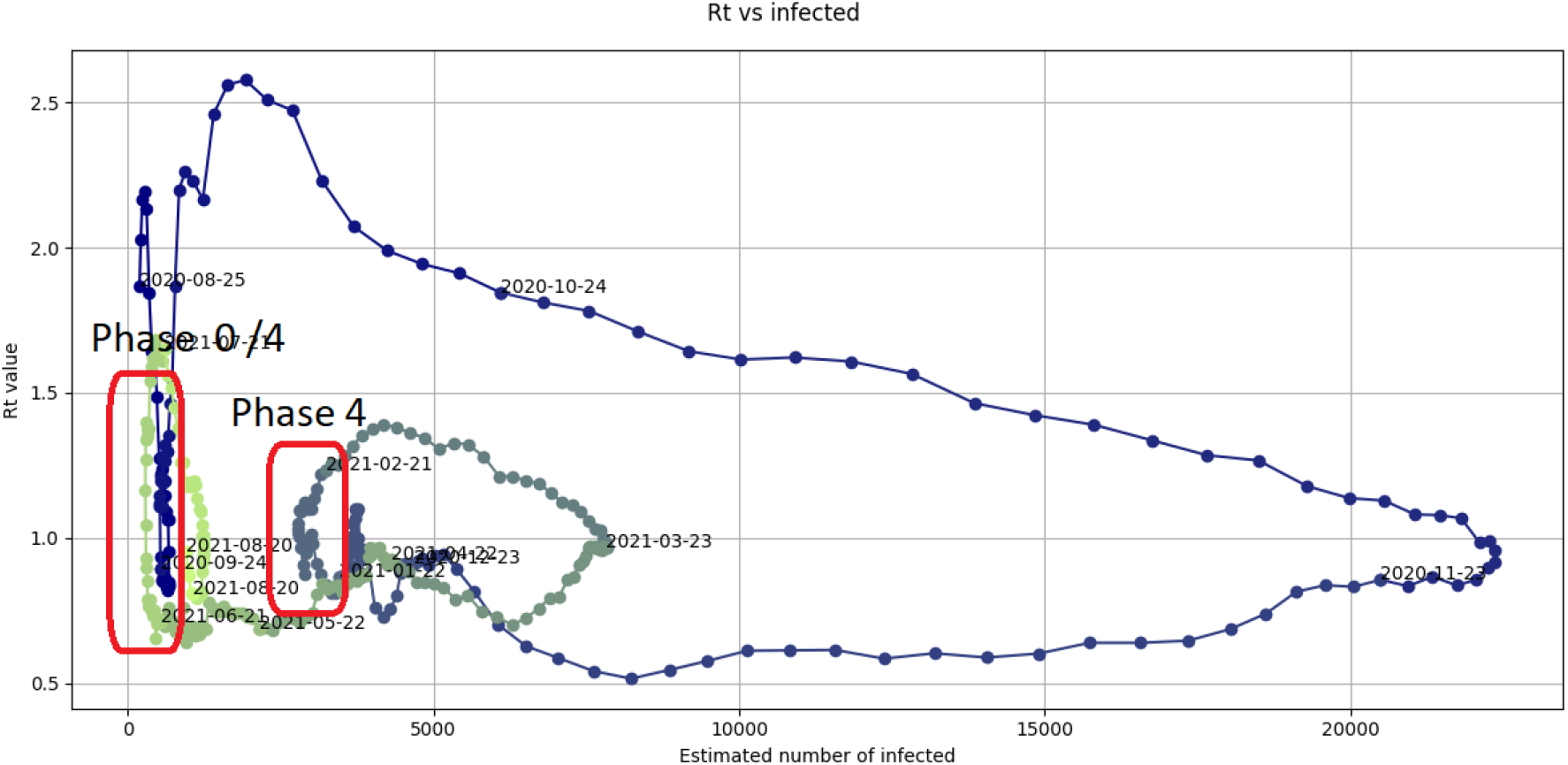
Graph of the values of the *R*_*t*_(*I*_*t*_) function observed in the province of Milan between August 25th, 2020 and August 24th, 2021. Phases 4 are highlighted.

**Figure 28:**
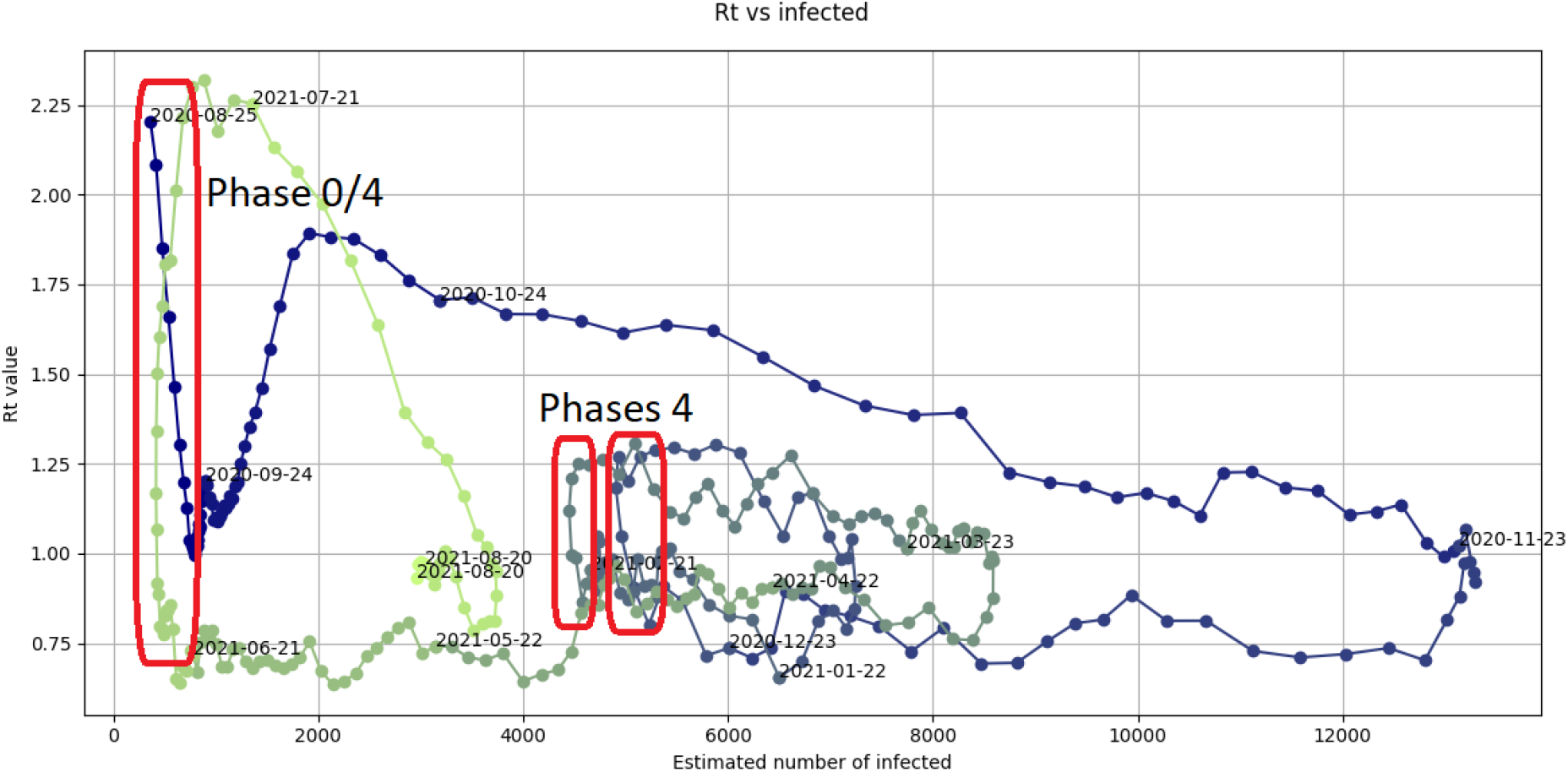
Graph of the values of the *R*_*t*_(*I*_*t*_) function observed in the province of Rome between August 25th, 2020 and August 24th, 2021. Phases 4 are highlighted.

**Figure 29:**
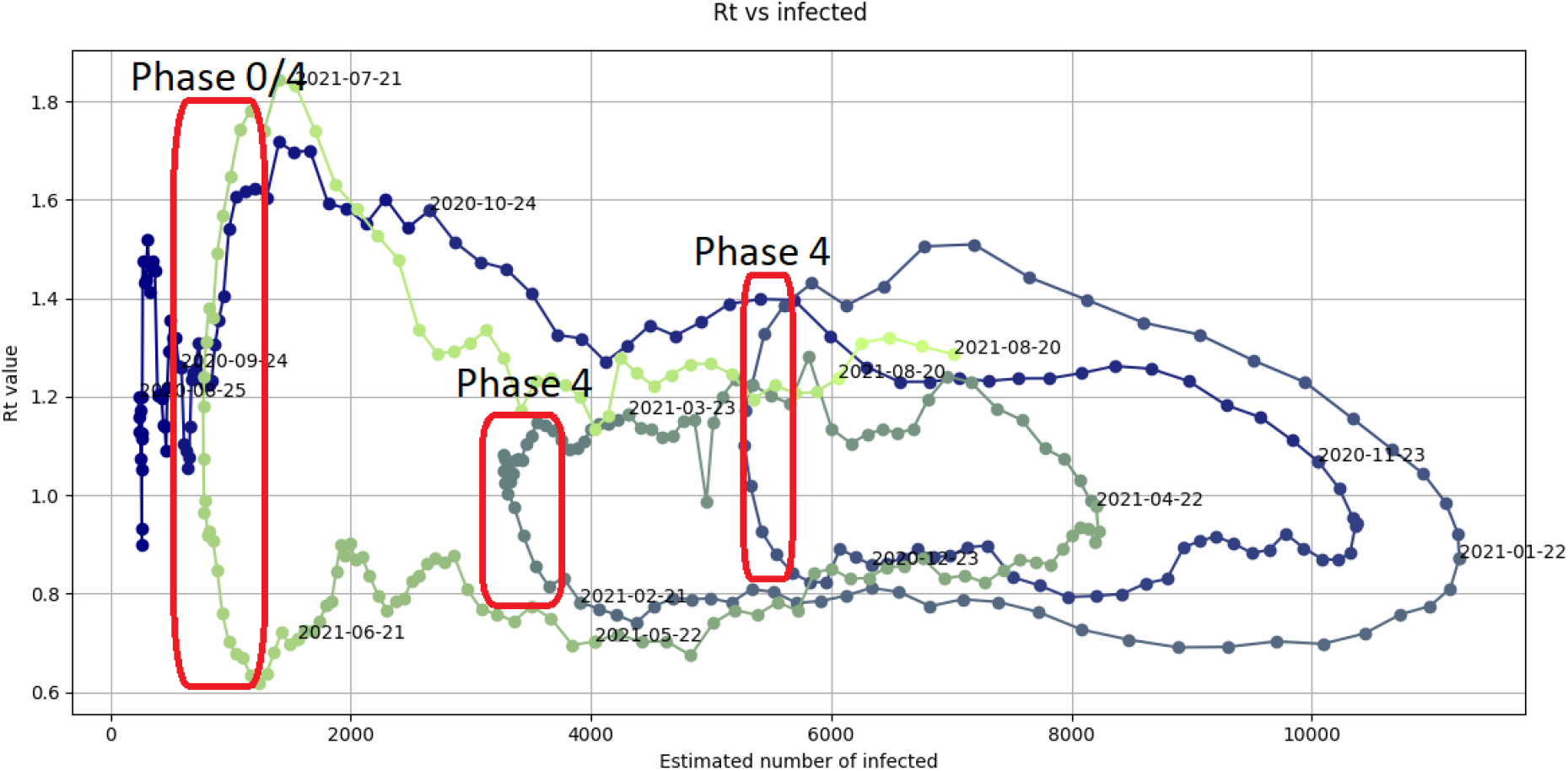
Graph of the values of the *R*_*t*_(*I*_*t*_) function observed in the region of Sicily between August 25th, 2020 and August 24th, 2021. Phases 4 are highlighted.

A Phase 4 corresponds to a new increase in contagions and, therefore, to the beginning of the rise of a new pandemic wave in the graph of the new infected individuals.

In Fig. 27-29, we can observe that there is not a clear cut that allow us to distinguish a Phase 4 from a Phase 0, when the number of infected individuals is relatively low. It can be also observed that a Phase 4 is always followed, possibly for a very short time, by a Phase 1. Pandemic waves apparently do not switch directly to a Phase 2.

Determining the reasons and the immediate detection of Phases 0 and 4 would allow the rapid implementation of containment measures to reduce the number and the size of future pandemic waves.

## 4 Conclusions

In this work we presented possible uses of the representation of the effective reproduction number *R*_*t*_ in the *I × R*_*t*_ space.

We showed that empirical data suggest that Phases 0, 2, and 4 have a brief duration and correspond to a clear modifications of the trend of contagions. An in-depth analysis of causes of these phases may allow us to identify which measures have a major effectiveness in containing the pandemic outbreak. Specifically, the study on how the temporal occurrence of the different phases in the various Italian regions and provinces can be traced back to national measures or to the seasonal trend of the pandemic may allow us to analyze which measures may appear more efficient to induce a Phase 2 or which critical issues arose over time that triggered a Phase 0 or 4. In particular, it is of interest to understand why the different phases may have occurred in different time periods in the different regions and provinces.

Given the typical trend of the pandemic, it might be worth trying to calculate a window where the ratio between *R*_*t*_ and infected individuals reaches the threshold necessary for a phase change. The knowledge of the values that characterize this window could lead to the immediate implementation of the measures that lead to the entry of Phase 2 of a loop, preventing the beginning of the new pandemic wave.

Finally, a word of caution must be spent. The immediate detection of the beginning of a Phase 0 or a Phase 4 in the graph of the *R*_*t*_(*I*_*t*_) function may be helpful in defeating the pandemic. Unfortunately, this operation is not obvious and false positive detections may occur.

## Data Availability

Data are openly available at https://github.com/pcm-dpc/COVID-19

http://virgo.unive.it/pesenti/tekwp/dashboard.php

